# Heart failure symptoms predict hospitalization and mortality at diagnosis, 6 and 12 month follow-ups

**DOI:** 10.1101/2024.06.12.24308679

**Authors:** MR Ali, CSP Lam, A Stromberg, SPP Hand, S Booth, F Zaccardi, GP McCann, K Khunti, CA Lawson

## Abstract

**Background:** We investigated symptoms reported before and after heart failure (HF) diagnosis and their associations with 3-month hospitalisation and mortality.

**Objectives:** To examine associations between symptoms recorded in primary care and short-term hospitalisation and mortality in HF patients.

**Design:** Landmark analysis using Royston-Parmar survival models at baseline (diagnosis), 6- and 12 months post-diagnosis.

**Setting:** Primary care database (CPRD) linked to hospital and mortality data (1998–2020).

**Participants:** Adults (>40 years) with a first HF diagnosis.

**Exposures:** Shortness of breath (SOB), ankle swelling, oedema, fatigue, chest pain, depression, and anxiety in the 3 months before diagnosis and at 6 and 12 months.

**Outcomes:** 3-month all-cause hospitalisation and mortality; secondary outcomes included HF and non-cardiovascular hospitalisation.

**Results:** Among 86,882 HF patients (62,742 and 54,555 surviving to 6 and 12 months, respectively), symptom associations varied by timepoint. At diagnosis, depression had the highest risk for all-cause hospitalisation (HR: 1.26; 95% CI 1.15, 1.39) and SOB for HF hospitalisation (1.18; 1.12, 1.26). At 6 months, depression was most associated with all-cause hospitalisation (1.46; 1.25, 1.70), and ankle swelling with mortality (1.49; 1.14, 1.94). At 12 months, SOB had the highest risk for HF hospitalisation (1.99; 1.68, 2.35).

**Conclusions:** Symptoms persisted and were more prominent at 6 and 12 months post-diagnosis than at diagnosis.

**Strengths and limitations:**

- This study used a large, nationally representative cohort from the Clinical Practice Research Datalink (CPRD), enhancing the generalisability of findings to the broader UK heart failure population.
- A dynamic prediction approach (landmark analysis) was employed to account for the time-varying and time-dependent nature of symptoms and covariates, addressing key limitations of static prognostic models.
- Routinely recorded primary care data were used to capture a broad range of HF-specific and non-specific symptoms, sociodemographic factors, treatments, and comorbidities across multiple clinically relevant timepoints.
- Symptoms and covariates were updated at each landmark, allowing for better reflection of patient status over time; however, landmark intervals were selected a priori and may not fully capture individual-level variation.
- Key clinical markers of heart failure severity (e.g., ejection fraction, NYHA class, natriuretic peptides) were not available, which may limit the assessment of symptom relevance across different HF phenotypes.

## INTRODUCTION

Heart failure (HF) is a complex clinical condition, often complicated by multiple long-term conditions (MLTC), that has inferior survival rates compared to numerous prevalent cancers.^1^ It affects over 64 million people worldwide^2,3^ with ever increasing prevalence.^4^ The global economic cost of HF had been estimated to be $108 billion^5^ amounting to 2% of annual health budgets.^6^ HF is the top cause of preventable hospitalisations in Europe^7^ and most of these costs are attributable to the high rate of hospitalisations,^8^ particularly for non-cardiovascular reasons.^9^ Given the 50% projected increase in HF hospitalisations by 2035, reducing hospitalisations in HF has become a key policy priority.^10,11^

Identifying individuals with chronic heart failure who are at risk of hospitalisation remains challenging. Existing prognostic models in HF primarily focus on mortality^12^ or are tailored for patients already admitted to the hospital,^13^ which limits their use in community settings. Moreover, existing models frequently rely on complex clinical data and biometrics measured at a single time point, exhibiting limited predictive efficacy.^14,15^

HF is a clinical diagnosis where patients present with symptoms typically before investigations have been undertaken.^16^ Individuals diagnosed with HF frequently encounter various indications and manifestations associated with fluid overload, such as breathlessness and ankle swelling.^16^ Additionally, they may experience other general symptoms that could be related to their HF condition or concurrent medical conditions, including fatigue, pain, and anxiety.^17^ Although symptoms are associated with hospitalisations and mortality,^18^ they are rarely used in prognosis.^19,20^

Symptoms in HF patients often fluctuate over time, meaning that their importance in prognosis may differ according to when they occur. For example, symptoms during the acute, diagnostic phase may have different associations with outcomes than those presenting later during the more chronic^20^ or end-stage.^21^ Furthermore, the association between symptoms and outcomes will likely depend on the follow-up period, with stronger associations in the short time after they are experienced. The time-varying and time-dependent nature of symptoms is rarely considered in risk prediction. Recently, the introduction of dynamic prediction, in particular landmark analysis, has provided a useful alternative to using a single-timepoint to estimate risk.^22^ Using this approach, symptoms and clinical characteristics are updated at different time-points and used to predict outcomes over a clinically relevant time horizon.^23^

In a UK national cohort of patients with a new diagnosis of HF, we investigated common symptoms reported prior to HF diagnosis and at 6 and 12-months post-diagnosis to identify their associations with 3-month all-cause and cause-specific hospitalisation and all-cause mortality.

## METHODS

### Study population

We used the Clinical Practice Research Datalink (CPRD), an anonymised electronic primary database covering over 11.3 million patients.^24^ This database is an internationally recognised population level database, representative of the general population in terms of age, sex, and ethnicity^24^ and includes information on sociodemographic, clinical, and lifestyle factors as well as laboratory and prescription data. CPRD uses a representative sample of general practices in the UK and is linked to national datasets including Hospital Episode Statistics (HES) and Office of National Statistics (ONS), providing hospital and mortality data, respectively.

Our study included individuals aged 18 years or older who had a first recorded diagnosis of heart failure (HF) in either their primary care or hospital records between 1^st^ January 1998 and 31^st^ March 2020. To be eligible for the study, patients needed to have at least one year of up-to-standard clinical data (a marker of the quality of data available in CPRD) available in CPRD prior to their inclusion and be eligible for linkage to hospital and death data. HF in primary care records was based on clinically validated terms,^25^ specifically focusing on Read codes (coded thesaurus of clinical terms used in the UK) within chapter G58, along with HF-specific Read codes from other chapters. For hospital records, we used ICD-10 codes for HF in the primary discharge position (Supplementary S1 Table for code lists). In cases where patients had both primary care and secondary care HF codes, the earliest recorded code as the HF index date, representing the date of diagnosis, was used.

### Exposure identification

To identify symptoms and signs pertinent to hospitalisation in heart failure (HF) patients, we employed patient consensus^26^ and national guidelines.^16^ These symptoms are routinely documented during clinical consultations in primary care. The selected symptoms encompassed shortness of breath, ankle swelling, general oedema (including both abdomen and peripheral oedema), fatigue, chest pain, general pain, anxiety, and depression. For each symptom, we utilised relevant Read codes that were recorded at least once in the three months prior to the landmark dates (diagnosis, 6, and 12 months). The code sets were reviewed and validated by a clinician (S2 Table).

### Measurement of covariates

Based on existing evidence,^27^ the covariates included in our analysis encompassed sociodemographic variables (age, sex, ethnicity, and socioeconomic status), lifestyle factors (body mass index (BMI), alcohol consumption, and smoking status), drug prescriptions (beta blockers, renin angiotensin system inhibitors (angiotensin-converting enzyme (ACE) inhibitors or angiotensin receptor blockers (ARB)), angiotensin receptor neprilysin inhibitors (ARNI), alpha blockers, sodium-glucose cotransporter (SGLT) 2 inhibitors, aspirin, statins, sodium-glucose cotransporter (SGLT) 2 inhibitors, and loop diuretics) and physiological data (systolic blood pressure, haemoglobin levels, cholesterol levels, and estimated glomerular filtration rate (eGFR)). Additionally, we accounted for several comorbidities, including hypertension, diabetes, ischaemic heart disease (IHD), myocarditis, cerebrovascular accident (stroke), atrial fibrillation (AF), asthma, cancer, cardiomyopathy, chronic obstructive pulmonary disease (COPD), chronic kidney disease (CKD), osteoarthritis (OA), rheumatoid arthritis (RA), dementia, and depression. As a surrogate measure for heart failure (HF) severity, we utilised hospitalisation history (within one month; one to three months; or three to 12 months prior to the landmark point).^28^

To assess socioeconomic status, we utilised the Index of Multiple Deprivation (IMD), a composite measure comprising seven indicators weighted to reflect deprivation in various domains such as income, employment, education, skills and training, health, crime, and barriers to housing.^29^ For chronic comorbidities, except for diabetes and depression, we considered the presence of at least one Read or ICD-10 code recorded before the landmark date. As depression may have intermittent occurrences, we defined it based on the presence of at least one code or prescription within a 12-month period prior to the landmark date. Diabetes was defined by clinical code or prescription at any time prior to the landmark date. Prescribed medications were identified through at least one prescription within the four months preceding the landmark date. All other covariates were captured based on their most recent values prior to the landmark date: these were updated at the different landmark points to account for changes in treatment status and disease history.

### Outcomes

Patients were followed up for 3-months after each landmark (diagnosis, and 6 and 12 month post diagnosis) point for first hospitalisation for any-cause, HF, and non-CVD causes; and for death.

### Statistical analysis

First, characteristics of included patients are presented for the different landmarks (diagnosis, 6, and 12 months after diagnosis). Second, at each landmark point, Royston Parmar flexible parametric survival models^30^ were used to estimate the association between each symptom and 3-month first all-cause and cause specific hospitalisation and mortality, using the “stpm2” command in Stata 17 (StataCorp, College Station, TX, USA). All individuals were included in the baseline model and all survivors at the subsequent landmarks in the 6 and 12- month models. Survivors were patients who were alive, had not transferred out of their practice and their practice was still contributing data to CPRD at the 6 and 12 month landmark dates. Assuming a missing at random mechanism, multiple imputation using chained equations was performed to impute socioeconomic status, ethnicity, and physiological data (missingness information is reported in Table 1), generating 10 imputed datasets with the results combined using Rubin’s rules.^31^ Multicollinearity between all variables in the model was assessed and linearity between each continuous variable and outcome was determined using likelihood ratio tests,^32^ Akaike and Bayesian Information Criteria (AIC and BIC), comparing models with different transformations (a quadratic extension or restricted cubic splines with three degrees of freedom). Unadjusted hazard ratios with 95% confidence intervals were estimated followed by adjustment of each symptom by all covariates. Depression comorbidity was not included in the 3-month history of depression symptom due to collinearity. A sensitivity analysis was undertaken using complete case analysis (S4 Table).

**Table 1:** Patient characteristics prior to and, 6-, and 12-months after HF diagnosis.

|  | Baseline<br>N=86,882 | 6 months<br>N=62,742 | 12 months<br>N=54,555 |
| --- | --- | --- | --- |
| Age at diagnosis (years) | 79.0 (71.0-86.0) | 78.0 (70.0-84.0) | 77.0 (69.0-84.0) |
| Female | 42,415 (49%) | 29,749 (47%) | 25,787 (47%) |
| **Ethnicity** |  |  |  |
| White | 77,442 (89%) | 57,404 (91%) | 50,215 (92%) |
| South Asian | 1,238 (1%) | 1,004 (2%) | 879 (2%) |
| Black | 637 (1%) | 520 (1%) | 437 (1%) |
| Other - mixed | 712 (1%) | 522 (1%) | 453 (1%) |
| Unknown | 4,638 (5%) | 2,350 (4%) | 1,860 (3%) |
| *Missing* | 2,215 (3%) | 942 (2%) | 711 (1%) |
| **IMD level** |  |  |  |
| 1 (least deprived) | 16,701 (19%) | 12,251 (20%) | 10,635 (19%) |
| 2 | 19,775 (23%) | 14,326 (23%) | 12,366 (23%) |
| 3 | 18,580 (21%) | 13,351 (21%) | 11,637 (21%) |
| 4 | 17,166 (20%) | 12,300 (20%) | 10,713 (20%) |
| 5 (most deprived) | 14,502 (17%) | 10,426 (17%) | 9,141 (17%) |
| *Missing* | 158 (<1%) | 88 (<1%) | 63 (<1%) |
| **HF diagnosis in secondary care** | 32,456 (37%) | 19,878 (32%) | 16,466 (30%) |
| **Previous any-cause hospitalisation** |  |  |  |
| None | 30,494 (35%) | 22,344 (36%) | 24,305 (45%) |
| Within 1 month | 44,738 (51%) | 3,714 (6%) | 2,883 (5%) |
| 1-3 months | 4,949 (6%) | 5,183 (8%) | 3,590 (7%) |
| 3-12 months | 6,701 (8%) | 31,501 (50%) | 23,777 (44%) |
| **Treatments** |  |  |  |
| RASI | 43,763 (50%) | 44,511 (71%) | 38,828 (71%) |
| ACE | 35,223 (41%) | 36,768 (59%) | 30,764 (56%) |
| Aspirin | 35,434 (41%) | 28,495 (45%) | 24,514 (45%) |
| Beta blocker | 28,665 (33%) | 28,320 (45%) | 24,990 (46%) |
| Statin | 31,428 (36%) | 28,069 (45%) | 25,271 (46%) |
| SGLT2 | 51 (<1%) | 49 (<1%) | 49 (<1%) |
| Loop diuretic | 45,987 (53%) | 45,292 (72%) | 37,631 (69%) |
| **Comorbidities** |  |  |  |
| AF | 34,513 (40%) | 27,065 (43%) | 24,068 (44%) |
| Cardiomyopathy | 3,190 (4%) | 3,881 (6%) | 3,909 (7%) |
| Hypertension | 56,013 (64%) | 42,110 (67%) | 37,277 (68%) |
| IHD | 43,922 (51%) | 34,058 (54%) | 30,641 (56%) |
| Myocarditis | 207 (<1%) | 202 (<1%) | 189 (<1%) |
| CVA | 16,090 (19%) | 11,389 (18%) | 9,992 (18%) |
| Diabetes | 20,783 (24%) | 15,764 (25%) | 13,354 (24%) |
| CKD (<60ml/min/m<sup>2</sup>) | 36,987 (56%) | 30,143 (59%) | 27,238 (59%) |
| Depression | 1,440 (2%) | 738 (1%) | 605 (1%) |
| Asthma | 16,562 (19%) | 12,578 (20%) | 11,195 (21%) |
| Cancer | 19,706 (23%) | 13,918 (22%) | 12,181 (22%) |
| COPD | 16,574 (19%) | 12,537 (20%) | 11,127 (20%) |
| Dementia | 4,199 (5%) | 2,499 (4%) | 2,127 (4%) |
| OA | 32,578 (37%) | 24,047 (38%) | 21,286 (39%) |
| RA | 6,354 (7%) | 4,618 (7%) | 4,099 (8%) |
| **Physiological/lifestyle** |  |  |  |
| BMI (Kg/m<sup>2</sup>) | 27.8 (±6.1) | 27.9 (±6.1) | 27.9 (±6.1) |
| eGFR (ml/min/m<sup>2</sup>) | 57.4 (±21.1) | 56.6 (±21.0) | 56.1 (±21.1) |
| Systolic BP (mmHg) | 137.7 ($\pm 22.0$) | 133.6 ($\pm 21.4$) | 132.8 ($\pm 20.5$) |
| Cholesterol (mmol/L) | 4.6 ($\pm 1.2$) | 4.6 ($\pm 1.2$) | 4.5 ($\pm 1.2$) |
| Haemoglobin (g/dL) | 12.9 ($\pm 1.9$) | 13.0 ($\pm 1.8$) | 13.0 ($\pm 1.8$) |
| **Smoking** |  |  |  |
| Yes | 17,317 (20%) | 11,687 (19%) | 9,852 (18%) |
| No | 38,386 (44%) | 27,482 (44%) | 23,791 (44%) |
| Ex | 25,722 (30%) | 20,460 (33%) | 18,525 (34%) |
| Missing | 5,457 (6%) | 3,113 (5%) | 2,387 (4%) |
| **Alcohol** |  |  |  |
| Yes | 52,753 (61%) | 39,248 (63%) | 34,407 (63%) |
| No | 18,554 (21%) | 13,318 (21%) | 11,624 (21%) |
| Ex | 2,973 (3%) | 2,323 (4%) | 2,148 (4%) |
| Missing | 12,602 (15%) | 7,853 (13%) | 6,376 (12%) |

Values are median (interquartile range), n (%), or mean ± SD. ACE = angiotensin-converting enzyme; ARB = angiotensin receptor blocker; ARNI = Angiotensin receptor-neprilysin inhibitor ; AF = atrial fibrillation; BMI = body mass index; BP = blood pressure; COPD = chronic obstructive pulmonary disease; CKD = chronic kidney disease; CVA = cerebrovascular accident (Stroke); eGFR = estimated glomerular filtration rate; Hb = haemoglobin; IHD = ischaemic heart disease; IMD = index multiple deprivation; OA = osteoarthritis; RA = rheumatoid arthritis; RASI = renin–angiotensin system inhibitor; SGLT2 = Sodium-glucose Cotransporter-2 inhibitors.

### Ethics approval

Approval for this study (protocol 20_000056) was provided by CPRD’s Research Data Governance Process. This process provides independent scientific and patient advice by an expert review and central advisory committee. National research ethics committee provided ethics approval for use of CPRD data (05/MRE04/87/AM06).

### Patient and public involvement

A consensus study^34^ was conducted with HF patients, carers, and their clinicians which identified a core set of symptoms experienced by people with HF before hospitalisation.

## RESULTS

At diagnosis (baseline) there were 86,882 patients with HF: the median age was 79 (IQR: 71- 86) years and 49% were female (Table 1). There were 62,742 survivors at 6 months and 54,555 (Figure 1) at 12-months (median age: 78 [70-84] and 77 [69-84] years, respectively; both 47% female). Prescription rates prior to diagnosis were relatively low (RASI, 50%; beta blocker, 33%) but increased in survivors by the 6 and 12 month landmark (71% and 45%, respectively). Prevalence of comorbidities were generally higher in survivors at 6 and 12- months whilst smoking reduced.

**Figure 1.**
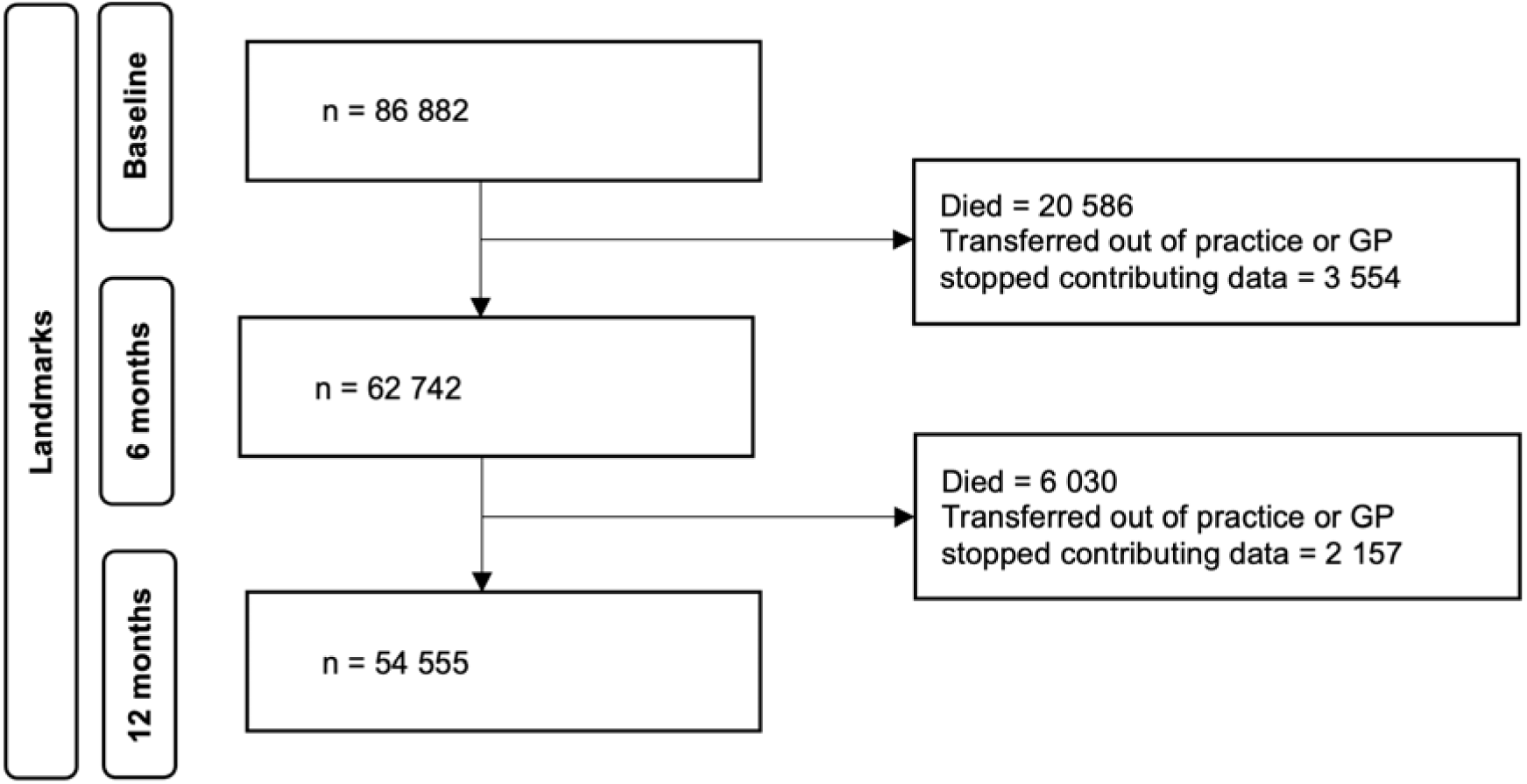
Flow chart of number of patients per landmark time

**Figure 2.**
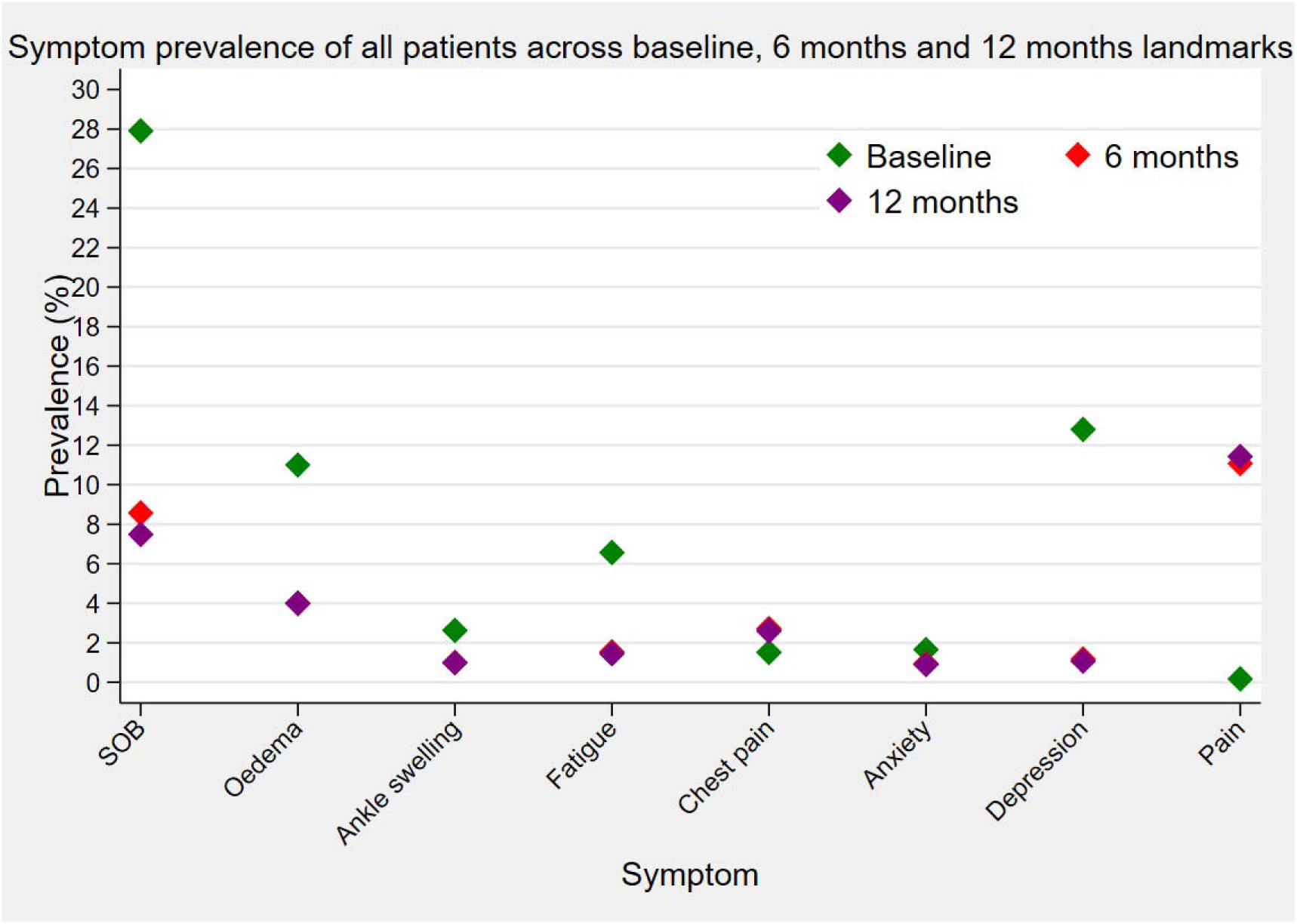
Symptom prevalence across baseline, 6-, and 12-months landmark times. Symptom prevalence were similar between 6 and 12 months landmark points so where a red diamond (denoted 6 month landmark) is not visible, it is behind the purple diamond (12 month landmark.

### Symptom prevalence across landmark times

Symptoms were measured in a three-month window prior to each landmark time (supplementary S3). The number of patients reporting at least one symptom was 42,327 (51.3%) prior to diagnosis; 15,613 (24.9%) prior to 6-month landmark; and 12,808 (23.5%) prior to 12-months. The most frequently recorded symptoms prior to diagnosis were SOB (27.9%), depression (12.8%), oedema (11.0%), and fatigue (6.6.%) (Figure 1). Pain was the most frequently recorded symptom prior to the 6- and 12-month landmarks (11.1% and 11.4%, respectively), followed by SOB (8.6% and 7.5%), oedema (both 4%), and chest pain (2.7% and 2.6%) (Figure 1).

### Symptom prediction when measured from diagnosis (baseline)

At baseline (symptoms measured prior to diagnosis), the symptoms associated with the highest risk of hospitalisation for any cause were depression (adjusted hazard ratio (aHR): 1.26; 95% confidence interval 1.15 to 1.39), fatigue (1.11; 1.02, 1.20) and chest pain (1.11; 1.05, 1.17) (Figure 3 and S3). For HF hospitalisation the symptoms associated with the highest risk were SOB (1.18; 1.12, 1.26) and general oedema (1.14; 1.05, 1.24), for non-cardiovascular admission were pain (1.15; 1.10, 1.21), depression (1.37; 1.22, 1.54) and fatigue (1.12; 1.02, 1.24) and for death were depression (1.22; 1.09, 1.36), and pain (1.06; 1.01, 1.11) (Figure 3). SOB showed a protective effect for the outcome of mortality (0.76; 0.74, 0.79) as did chest pain (0.79; 0.74, 0.85) (Figure 4). Complete case-analyses for all outcomes can be found in S4.

**Figure 3.**
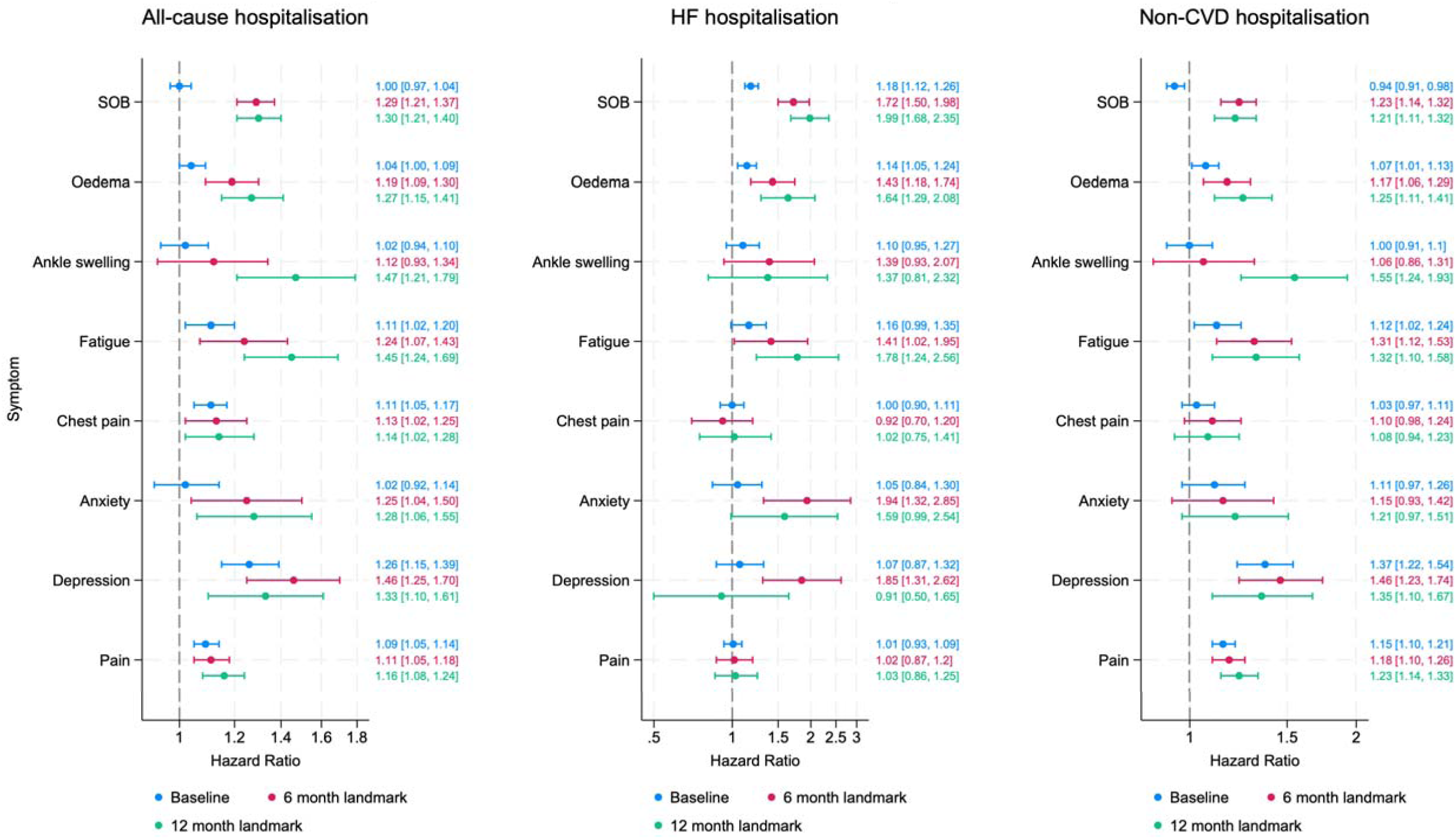
Symptom associations across three different landmarks for all-cause hospitalisation, HF and non-CVD hospitalisation Forest plot of adjusted effect estimates between symptoms and hospitalisation. Survival models at three different landmarks adjusted for the following covariates: age, sex, ethnicity, deprivation, BMI, diagnosis in hospital or primary care, comorbidities and depression. BMI, body mass index; CVD, cardiovascular disease; HF, heart failure.

**Figure 4.**
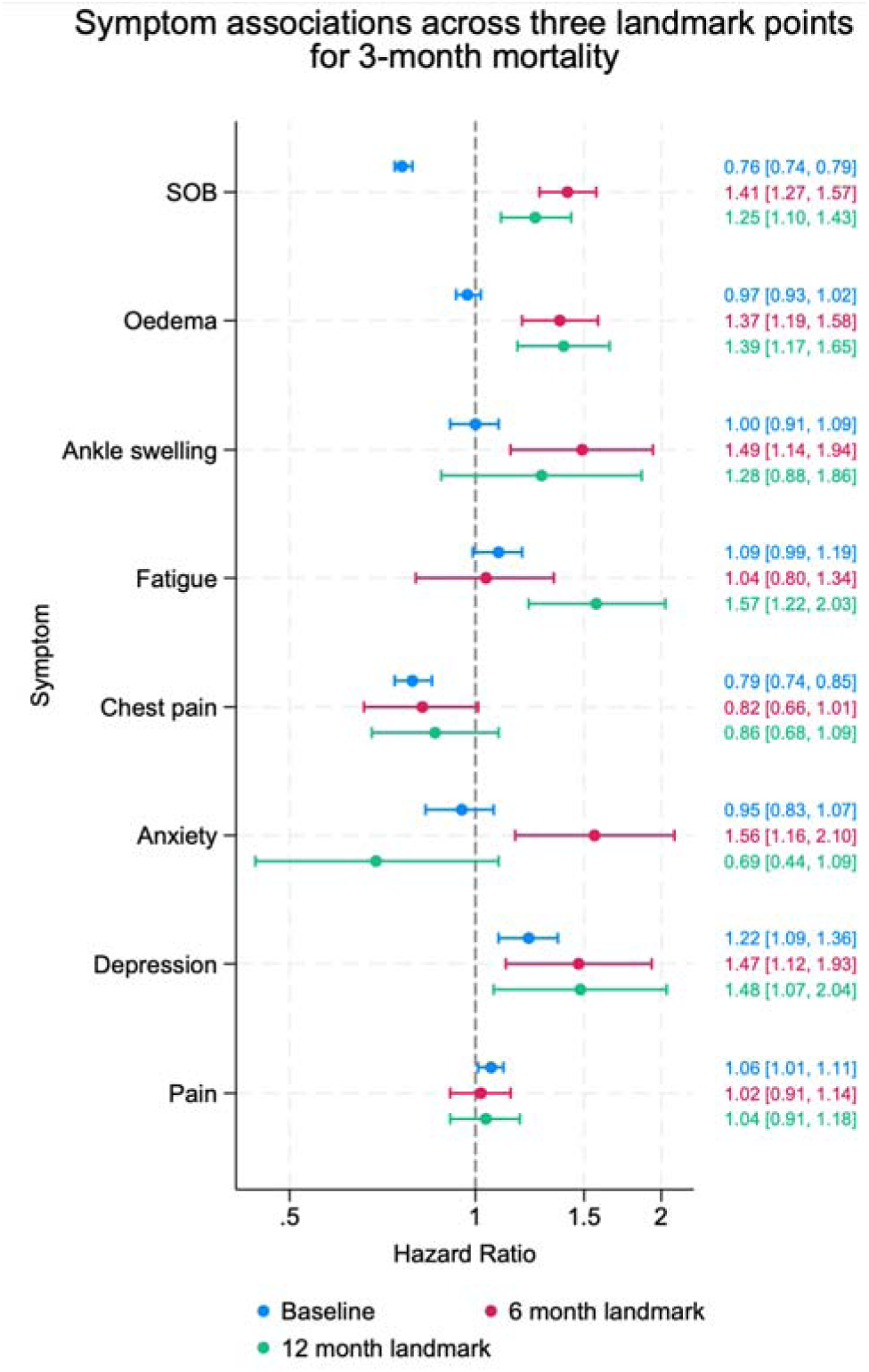
Symptom associations across three different landmarks for all-cause mortality Forest plot of adjusted effect estimates between symptoms and hospitalisation. Flexible parametric survival models at three different landmarks adjusted for the following covariates: age, sex, ethnicity, deprivation, BMI, diagnosis in hospital or primary care, comorbidities and depression. BMI, body mass index.

### Symptom prediction when measured from 6-month landmark time

Whilst the prevalence of recorded symptoms was lower at the 6 and 12-months landmarks, the strength of associations between symptoms and all outcomes generally increased. At the 6-month landmark time, the symptoms associated with the highest risk of hospitalisation for any cause were depression (aHR: 1.46; 95% CI: 1.25, 1.70), SOB (1.29; 1.21, 1.37), anxiety (1.25; 1.04, 1.50), fatigue (1.24; 1.07, 1.43), general oedema (1.19; 1.09, 1.30), chest pain (1.13; 1.02, 1.25) and general pain (1.11; 1.05, 1.18); for HF admission were anxiety (1.94; 1.32, 2.85), depression (1.85; 1.31, 2.62), SOB (1.72; 1.50, 2.98), general oedema (1.43; 1.18, 1.74) and fatigue (1.41; 1.02, 1.95); and for non-CVD admission were depression (1.46; 1.23, 1.74), fatigue (1.31; 1.12, 1.53), SOB (1.23; 1.14, 1.32), general pain (1.18; 1.10, 1.26), and general oedema (1.17; 1.06, 1.29) (Figure 3). Finally, for mortality, the symptoms associated with the highest risk were anxiety (1.56; 1.16, 2.10), ankle swelling (1.49; 1.14, 1.94), depression (1.47; 1.12, 1.93), SOB (1.41; 1.27, 1.57) and general oedema (1.37; 1.19, 1.58) (Figure 4).

### Symptom prediction when measured from 12-month landmark time

From the 12-months landmark, all the symptoms had statistically significant risk associations with any cause of hospitalisation. The symptoms associated with the highest risk of hospitalisation for any cause were ankle swelling (aHR 1.47; 95% CI: 1.21, 1.79), fatigue (1.45; 1.24, 1.69) and depression (1.33; 1.10, 1.61); for HF admission were SOB (1.99; 1.68, 2.35), fatigue (1.78; 1.24, 2.56) and general oedema (1.64; 1.29, 2.08); and for non-CVD admission they were ankle swelling (1.55; 1.24, 1.93), depression (1.35; 1.10, 1.67), fatigue (1.32; 1.10, 1.58), general oedema (1.25; 1.11, 1.41), general pain (1.23; 1.14, 1.33) and SOB (1.21; 1.11, 1.32) (Figure 3). Symptoms associated with the highest risk of mortality were fatigue (1.57; 1.22, 2.03), depression (1.48; 1.07, 2.04), oedema (1.39; 1.17, 1.65). Compared to the baseline where SOB was protective, at 12 months it showed a significant and increased risk of mortality (1.25; 1.10, 1.43) (Figure 4).

## DISCUSSION

The timing of symptom recording in relation to the outcome is crucial, especially considering the transient nature of symptoms.^33,34^ To our knowledge, this is the first study in the general practice population of patients with HF to report symptom risk associations, accounting for their time-varying and time-dependent nature. We found HF specific and generic symptoms to be significantly associated with increased rates of 3-month hospitalisation and mortality at all three timepoints, with stronger associations in a more stable state population at 6 and 12- months. Furthermore, symptoms recorded in routine primary care were found to modestly improve the discrimination of prognostic models, with good model performance at 6 and 12- months. This has important clinical significance for identifying patients with HF in the community, who are at the highest risk of imminent hospitalisation and mortality.

Prior evidence has shown conflicting evidence in the risk associations between symptoms and clinically important outcomes such as hospitalisation and mortality in HF.^35–38^ However, this may be explained by studies using a single time-point, usually at diagnosis or during hospitalisation (where patients are often less stable), to measure symptoms. Failure to account for the time-varying nature of symptoms may contribute to the conflicting evidence as changes in treatments, severity of disease, and newer diagnoses of comorbidities are likely to confound these associations. By using landmark analysis, we were able to account for changes in symptoms and important covariates such as prescribed treatments and physiological status.

Psychological symptoms, such as depression and anxiety, are important predictors in HF and yet they continue to be under recognised in prognosis.^39^ With the exception of HF admission, depression prior to diagnosis was a key predictor of all outcomes. Anxiety and depression were also key symptoms at 6 months, with anxiety associated with a 2-fold increase and depression an 80% increase in rates of hospitalisation for HF. Anxiety at 6 months was also associated with a 50% increase in the mortality rate. Consistent with previous recommendations, routine assessments of anxiety and depression symptoms in the HF patient population are urgently required.^40–42^

HF and non-HF symptoms are important indicators of clinical status and used in routine assessment,^16^ but little is known about their prognostic importance, with conflicting evidence in prediction.^35–38^ Notably, the risk associations between non-HF symptoms and outcomes (including HF admission) merits attention, especially as guidelines focus on HF specific symptoms.^16^ This study underscores the importance of both HF and non-HF symptoms in risk assessments.

Most admissions in HF patients are for non-CVD causes. Our study revealed robust and higher risk associations between HF symptoms and non-CVD hospitalisation, implying HF patients may be admitted to hospital for non-CVD causes that could be due to their MLTCs. This is particularly concerning given that HF patients to non-cardiology wards have different demographics compared to those treated in cardiology services and have higher risk of adverse outcomes compared to patients admitted to cardiology wards.^43^

This study shows that symptoms could be an important and early indicator of imminent hospital admission. However, there is an urgent need to develop ways for patients to monitor these symptoms more closely. Many patients rarely formally record their symptoms and some don’t report them. Despite the mounting pressures on consultation times in primary care,^44^ utilising symptoms at diagnosis and beyond emerges as a potentially simple and cost-effective method to identify high risk patients, compared to using blood tests or expensive scans, where there are delays in provision. This is particularly important as a growing body of research has shown PROMs recording tools such as Kansas City Cardiomyopathy Questionnaires (KCCQ) able to predict imminent deterioration and risk of hospitalisation and mortality.^20,45^ Albeit, an important consideration is the extra time and resource required to record PROMs and any implementation should be carried out in consultation with primary care health professional and patients.^46^ That said, challenges persist for patients and clinicians in recognising relevant symptoms,^47^ particularly patients with a multimorbid history.^48^ Overall, symptoms may prove to be a valuable, easily accessible method for improving patient outcomes and should be considered and prioritised in future risk prediction strategies.

Our data showed a variation in the associations between symptoms and outcomes at different timepoints. Symptoms recorded 6 months and 12 months post-diagnosis had stronger associations with outcomes than those observed at diagnosis. Given the high mortality risk in the first few months following HF diagnosis,^16,28^ this variation is likely due to the lower baseline risk in survivors at 6-months and beyond, leading to higher relative effects and better discrimination and highlights the importance of symptom monitoring in the community where most HF patients are managed.

The study strengths include the large national database of people with a new diagnosis of HF over 20-years. To account for the high comorbidity burden in HF patients and the variations in symptom presentation, we used a broad spectrum of symptoms recorded routinely in primary care at different timepoints. Using the landmark analysis approach, we were able to account for the time-varying nature of symptoms and covariates and investigate the temporal relationship between symptoms and imminent hospitalisation and mortality.

However, several limitations need to be acknowledged. Although there is some evidence of large population databases in primary care recording symptoms,^49^ these may be incomplete. However, under reporting would likely bias the associations towards the null value, leading to underestimated associations in this study. We did not have access to HF phenotype information and further work is needed to ascertain whether symptom associations differ by phenotype. Likewise, we did not have HF severity measures such as ejection fraction, NYHA, or natriuretic peptides. Moreover, we chose the landmarks a priori for clinical reasons. Shorter, longer, or more frequent landmark times may provide better data on important timepoints for symptoms and requires further research.

In conclusion, symptoms recorded routinely in primary care were significantly associated with increased rates of hospitalisation and death and provide the potential for a simple and patient centred approach to prognosis. Of particular clinical note is our findings that symptoms in patients with HF 6 and 12 month post diagnosis may be more important clinically and prognostically than those experienced around their diagnosis date. Efforts to improve the reporting and recording of symptoms are urgently required.

## SUMMARY BOX

### What is already known on this topic

- Heart failure (HF) is the leading cause of preventable hospitalisations in Europe.
- The identification of imminent risk of hospitalisation in HF poses a critical challenge and existing prediction models utilise complex information, lack patient-centeredness, and demonstrate suboptimal performance.
- Symptoms and other biomarkers are often recorded at a single time-point to predict risk without accounting for the time-varying and time-dependent nature of symptoms.

### What this study adds

- Common symptoms among HF patients are associated with a significant risk of both imminent hospitalisations and mortality.
- The presence of both HF-specific and general symptoms plays a crucial role in predicting the risk of HF-related hospitalisations, emphasising the significance of integrated care for heart failure and comorbidities.
- Considering symptoms as time-varying covariates is important clinically as we have shown symptoms recorded post diagnosis can be even more important than those reported at diagnosis.

### Data sharing

MRA and CL had full access to all the data in the study and takes responsibility for the integrity of the data and the accuracy of the data analysis. This study is based in part on data from the Clinical Practice Research Datalink obtained under licence from the UK Medicines and Healthcare products Regulatory Agency. However, the interpretation and conclusions contained in this report are those of the author/s alone. Data access is through permissions from CPRD only.

## Supporting information

STROBE checklist

## Acknowledgements

KK is supported by the National Institute for Health Research (NIHR) Applied Research Collaboration East Midlands (ARC EM) and the NIHR Leicester Biomedical Research Centre (BRC). GPM is funded by the NIHR (RP-2017-08-ST2-007) and receives support from the NIHR Leicester BRC. This research used the ALICE High Performance Computing Facility at the University of Leicester.

## Transparency statement

The lead author (MRA) affirms that the manuscript is an honest, accurate, and transparent account of the study being reported; that no important aspects of the study have been omitted; and that any discrepancies from the study as originally planned have been explained.

## Contributors

MRA— guarantor of the research, literature search, study design, data analysis, data interpretation, figures and writing. CSPL—data interpretation and writing. AS—data interpretation and writing. SPPH—figures, data interpretation and writing. SB—figures, study design, data interpretation and writing. FZ—study design, data interpretation and writing. GPM—data interpretation and writing. KK—study design, data interpretation and writing. CAL—literature search, study design, data analysis, data interpretation, figures and writing.

## Transparency

The lead author affirms that the manuscript is an honest, accurate, and transparent account of the study being reported, that no important aspects of the study have been omitted, and that any discrepancies from the study as originally planned (and, if relevant, registered) have been explained.

## Patient and public involvement

We worked in full collaboration with patients to design the study. We conducted a detailed consensus study^26^ with people with HF, their carers and HF clinicians, to identify a core set of symptoms and signs that are important to patients prior to hospitalisation. These symptoms and signs were used to design the exposures for this study.

