## Supplementary material for "Heart failure symptoms predict hospitalization and mortality at diagnosis, 6 and 12 month follow-ups": STROBE checklist

STROBE Statement—checklist of items that should be included in reports of observational studies

Study: **Association between routinely reported symptoms and signs in people with heart failure and 3-month hospitalisation and mortality: a study in the UK general population of >85,000 heart failure patients.**

|  | Item No. | Recommendation | Page  No. | Relevant text from manuscript |
| --- | --- | --- | --- | --- |
| **Title and abstract** | 1 | (*a*) Indicate the study’s design with a commonly used term in the title or the abstract | 2 | Landmark modelling. |
|  |  | (*b*) Provide in the abstract an informative and balanced summary of what was done and what was found | 2 |  |
| Introduction | | | |  |
| Background/rationale | 2 | Explain the scientific background and rationale for the investigation being reported | 4-5 | Given the 50% projected increase in HF hospitalisations by 2035, reducing hospitalisations in HF has become a key policy priority.^10,11^ However, identifying people with chronic heart failure who are at risk of imminent hospitalisation is a critical challenge. Symptoms in HF patients often fluctuate over time, meaning that their importance in prognosis may differ according to when they occur. For example, symptoms during the acute, diagnostic phase may have different associations with outcomes than those presenting later during the more chronic^20^ or end-stage.^21^ Furthermore, the association between symptoms and outcomes will likely depend on the follow-up period, with stronger associations in the short time after they are experienced. The time-varying and time-dependent nature of symptoms is rarely considered in risk prediction. Recently, the introduction of dynamic prediction, in particular landmark analysis, has provided a useful alternative to using a single-timepoint to estimate risk.^22^ Using this approach, symptoms and clinical characteristics are updated at different time-points and used to predict outcomes over a clinically relevant time horizon.^23^  In a UK national cohort of patients with a new diagnosis of HF, we investigated common symptoms reported prior to HF diagnosis and at 6 and 12-months post-diagnosis to identify their associations with 3-month all-cause and cause-specific hospitalisation and all-cause mortality. |
| Objectives | 3 | State specific objectives, including any pre specified hypotheses | 5 and 8 | In a UK national cohort of patients with a new diagnosis of HF, we investigated common symptoms reported prior to HF diagnosis and at 6 and 12-months post-diagnosis to identify their associations with 3-month all-cause and cause-specific hospitalisation and all-cause mortality. First, characteristics of included patients are presented for the different landmarks (diagnosis, 6, and 12 months after diagnosis). Second, at each landmark point, Royston Parmar flexible parametric survival models^30^ were used to estimate the association between each symptom and 3-month first all-cause and cause specific hospitalisation and mortality, using the “stpm2” command in Stata 17 (StataCorp, College Station, TX, USA). |
| Methods | | | |  |
| Study design | 4 | Present key elements of study design early in the paper | 6 | Our study included individuals aged 18 years or older who had a first recorded diagnosis of heart failure (HF) in either their primary care or hospital records between 1^st^ January 1998 and 31^st^ March 2020. To be eligible for the study, patients needed to have at least one year of up-to-standard clinical data (a marker of the quality of data available in CPRD) available in CPRD prior to their inclusion and be eligible for linkage to hospital and death data. |
| Setting | 5 | Describe the setting, locations, and relevant dates, including periods of recruitment, exposure, follow-up, and data collection | 6 | We used the Clinical Practice Research Datalink (CPRD), an anonymised electronic primary database covering over 11.3 million patients.^24^ This database is an internationally recognised population level database, representative of the general population in terms of age, sex, and ethnicity^24^ and includes information on sociodemographic, clinical, and lifestyle factors as well as laboratory and prescription data. CPRD uses a representative sample of general practices in the UK and is linked to national datasets including Hospital Episode Statistics (HES) and Office of National Statistics (ONS), providing hospital and mortality data, respectively. |
| Participants | 6 | (*a*) *Cohort study*—Give the eligibility criteria, and the sources and methods of selection of participants. Describe methods of follow-up  *Case-control study*—Give the eligibility criteria, and the sources and methods of case ascertainment and control selection. Give the rationale for the choice of cases and controls  *Cross-sectional study*—Give the eligibility criteria, and the sources and methods of selection of participants | 6-9  S1 | Our study included individuals aged 18 years or older who had a first recorded diagnosis of heart failure (HF) in either their primary care or hospital records between 1st January 1998 and 31st March 2020. To be eligible for the study, patients needed to have at least one year of up-to-standard clinical data (a marker of the quality of data available in CPRD) available in CPRD prior to their inclusion and be eligible for linkage to hospital and death data. HF in primary care records was based on clinically validated terms,25 specifically focusing on Read codes (coded thesaurus of clinical terms used in the UK) within chapter G58, along with HF-specific Read codes from other chapters. For hospital records, we used ICD-10 codes for HF in the primary discharge position (Supplementary S1 Table for code lists). In cases where patients had both primary care and secondary care HF codes, the earliest recorded code as the HF index date, representing the date of diagnosis, was used.  First, characteristics of included patients are presented for the different landmarks (diagnosis, 6, and 12 months after diagnosis). Second, at each landmark point, Royston Parmar flexible parametric survival models^30^ were used to estimate the association between each symptom and 3-month first all-cause and cause specific hospitalisation and mortality |
|  |  | (*b*) *Cohort study*—For matched studies, give matching criteria and number of exposed and unexposed  *Case-control study*—For matched studies, give matching criteria and the number of controls per case |  |  |
| Variables | 7 | Clearly define all outcomes, exposures, predictors, potential confounders, and effect modifiers. Give diagnostic criteria, if applicable | 6-8  S1 and S2 Table | Definition of outcomes, exposures and covariates. |
| Data sources/ measurement | 8* | For each variable of interest, give sources of data and details of methods of assessment (measurement). Describe comparability of assessment methods if there is more than one group | 6-7  S2 Table | Symptom exposures  Measurement of covariates |
| Bias | 9 | Describe any efforts to address potential sources of bias | 6  6  8  8 | Linkage of primary care and hospital data to reduce bias in the ascertainment of incident HF  Exclusion of people without at least one year of clinical data to reduce ascertainment bias.  Measurement of exposures and covariates prior to the match date  Multiple imputation & sensitivity analyses using full case analysis |
| Study size | 10 | Explain how the study size was arrived at | 6 | We included all patients aged ≥ 18 years, with a first code for HF within their primary care or hospital record between 1st January 1998 and 31st March 2020 |

Continued on next page

| Quantitative variables | 11 | Explain how quantitative variables were handled in the analyses. If applicable, describe which groupings were chosen and why | 9 | Multicollinearity between all variables in the model was assessed and linearity between each continuous variable and outcome was determined using likelihood ratio tests,32 Akaike and Bayesian Information Criteria (AIC and BIC), comparing models with different transformations (a quadratic extension or restricted cubic splines with three degrees of freedom). |
| --- | --- | --- | --- | --- |
| Statistical methods | 12 | (*a*) Describe all statistical methods, including those used to control for confounding | 8-9 | …at each landmark point, Royston Parmar flexible parametric survival models30 were used to estimate the association between each symptom and 3-month first all-cause and cause specific hospitalisation and mortality, using the “stpm2” command in Stata 17 (StataCorp, College Station, TX, USA). All individuals were included in the baseline model and all survivors at the subsequent landmarks in the 6 and 12-month models. Survivors were patients who were alive, had not transferred out of their practice and their practice was still contributing data to CPRD at the 6 and 12 month landmark dates. |
|  |  | (*b*) Describe any methods used to examine subgroups and interactions | 8 | With sub grouped admissions into subgroups by cause |
|  |  | (*c*) Explain how missing data were addressed | 9 | Multiple imputation using chained equations was performed (assuming a missing at random mechanism), to reduce the bias from missing socioeconomic status, ethnicity and physiological data and results were combined using Rubin’s rules.^38^ |
|  |  | (*d*) *Cohort study*—If applicable, explain how loss to follow-up was addressed  *Case-control study*—If applicable, explain how matching of cases and controls was addressed  *Cross-sectional study*—If applicable, describe analytical methods taking account of sampling strategy |  |  |
|  |  | (*e*) Describe any sensitivity analyses | 9 (S4 table) | A sensitivity analysis was undertaken using complete case analysis. |
| Results | | | | |
| Participants | 13* | (a) Report numbers of individuals at each stage of study—eg numbers potentially eligible, examined for eligibility, confirmed eligible, included in the study, completing follow-up, and analysed | 10-11  Table 1 | At diagnosis (baseline) there were 86,882 patients with HF: the median age was 79 (IQR: 71-86) years and 49% were female (Table 1). There were 62,742 survivors at 6 months and 54,555 (Figure 1) at 12-months (median age: 78 [70-84] and 77 [69-84] years, respectively; both 47% female). |
|  |  | (b) Give reasons for non-participation at each stage |  | N/A |
|  |  | (c) Consider use of a flow diagram | Figure 1 |  |
| Descriptive data | 14* | (a) Give characteristics of study participants (eg demographic, clinical, social) and information on exposures and potential confounders | Table 1 |  |
|  |  | (b) Indicate number of participants with missing data for each variable of interest | Table 1 | All missing data reported |
|  |  | (c) *Cohort study*—Summarise follow-up time (eg, average and total amount) | N/A |  |
| Outcome data | 15* | *Cohort study*—Report numbers of outcome events or summary measures over time | 10-11 | Numbers and percentages given |
|  |  | *Case-control study—*Report numbers in each exposure category, or summary measures of exposure |  |  |
|  |  | *Cross-sectional study—*Report numbers of outcome events or summary measures |  | N/A |
| Main results | 16 | (*a*) Give unadjusted estimates and, if applicable, confounder-adjusted estimates and their precision (eg, 95% confidence interval). Make clear which confounders were adjusted for and why they were included | Figures 3 &4  S3 Table | Unadjusted hazard ratios with 95% confidence intervals were estimated followed by adjustment of each symptom by all covariates. |
|  |  | (*b*) Report category boundaries when continuous variables were categorized |  | N/A |
|  |  | (*c*) If relevant, consider translating estimates of relative risk into absolute risk for a meaningful time period |  | N/A |

| Other analyses | 17 | Report other analyses done—eg analyses of subgroups and interactions, and sensitivity analyses | Figures 1 & 2 and S3 Table | Estimates presented for cause specific admissions subgroup |
| --- | --- | --- | --- | --- |
| Discussion | | | | |
| Key results | 18 | Summarise key results with reference to study objectives | 12 | Discussion, paragraph 1 |
| Limitations | 19 | Discuss limitations of the study, taking into account sources of potential bias or imprecision. Discuss both direction and magnitude of any potential bias | 12 | Study limitations |
| Interpretation | 20 | Give a cautious overall interpretation of results considering objectives, limitations, multiplicity of analyses, results from similar studies, and other relevant evidence | 11-13 | Discussion |
| Generalisability | 21 | Discuss the generalisability (external validity) of the study results | 13 | Further research required |
| Other information | |  | | |
| Funding | 22 | Give the source of funding and the role of the funders for the present study and, if applicable, for the original study on which the present article is based | 14 | The research was supported by an NIHR Advanced Fellowship [Reference NIHR300111]. |

**Note:** An Explanation and Elaboration article discusses each checklist item and gives methodological background and published examples of transparent reporting. The STROBE checklist is best used in conjunction with this article (freely available on the Web sites of PLoS Medicine at http://www.plosmedicine.org/, Annals of Internal Medicine at http://www.annals.org/, and Epidemiology at http://www.epidem.com/). Information on the STROBE Initiative is available at www.strobe-statement.org.
