## Supplementary data for "Heart failure symptoms predict hospitalization and mortality at diagnosis, 6 and 12 month follow-ups"

**Supplementary file**

**Association between routinely reported symptoms and imminent hospital admission and mortality: a landmark analysis study in a UK general population of >86,000 heart failure patients**

Ali MR^a,b,c^, Lam CSP^d,e^, Stromberg A^f,g^, Hand SPP^h^, Booth S^h^, Zaccardi F^c,i^, McCann GP ^a,b^, Khunti K^c,i^, Lawson CA^a,b,c^

**Affiliations**

a. Department of Cardiovascular Sciences, University of Leicester, UK

b. NIHR Leicester Cardiovascular Biomedical Research Unit, Glenfield Hospital, Leicester, UK

c. Leicester Real World Evidence Unit, University of Leicester, UK

d. National Heart Centre Singapore, 5 Hospital Drive, Singapore.

e. Duke-National University of Singapore Medical School, Singapore.

f. Department of Health, Medicine and Caring Sciences, Linköping University, Linköping, Sweden.

g. Department of Cardiology, Linköping University, Linköping, Sweden.

h. Department of Population Health Sciences, University of Leicester

i. Diabetes Research Centre, University of Leicester, UK

### **S1 Table: Read codes and ICD-10 code list for heart failure**

| readcode | readterm | medcode |
| --- | --- | --- |
| 1O1..00 | Heart failure confirmed | 9913 |
| 8H2S.00 | Admit heart failure emergency | 32898 |
| G1yz100 | Rheumatic left ventricular failure | 22262 |
| G232.00 | Hypertensive heart&renal dis wth (congestive) heart failure | 21837 |
| G234.00 | Hyperten heart&renal dis+both(congestv)heart and renal fail | 57987 |
| G58..00 | Heart failure | 2062 |
| G58..11 | Cardiac failure | 1223 |
| G580.00 | Congestive heart failure | 398 |
| G580.11 | Congestive cardiac failure | 2906 |
| G580.12 | Right heart failure | 10079 |
| G580.13 | Right ventricular failure | 10154 |
| G580.14 | Biventricular failure | 9524 |
| G580000 | Acute congestive heart failure | 23707 |
| G580100 | Chronic congestive heart failure | 32671 |
| G580200 | Decompensated cardiac failure | 27884 |
| G580300 | Compensated cardiac failure | 11424 |
| G580400 | Congestive heart failure due to valvular disease | 94870 |
| G581.00 | Left ventricular failure | 884 |
| G581.11 | Asthma - cardiac | 23481 |
| G581.12 | Pulmonary oedema - acute | 43618 |
| G581.13 | Impaired left ventricular function | 5942 |
| G581000 | Acute left ventricular failure | 5255 |
| G582.00 | Acute heart failure | 27964 |
| G583.00 | Heart failure with normal ejection fraction | 101138 |
| G583.11 | HFNEF - heart failure with normal ejection fraction | 101137 |
| G583.12 | Heart failure with preserved ejection fraction | 106897 |
| G584.00 | Right ventricular failure | 104275 |
| G58z.00 | Heart failure NOS | 4024 |
| G58z.11 | Weak heart | 12590 |
| G58z.12 | Cardiac failure NOS | 17278 |
| ICD-10 code | **Term** |  |
| I50 | Heart failure |  |
| 150.0 | Congestive heart failure |  |
| I50.1 | Left Ventricular failure |  |
| I50.9 | Heart failure, unspecified |  |
| I11.0 | Hypertensive heart disease with (congestive) heart failure |  |
| I13.2 | Hypertensive heart and renal disease with both (congestive) heart failure and renal failure |  |
| I13.0 | Hypertensive heart and renal disease with (congestive) heart failure |  |

### **S2 Table: Read code list for symptoms**

| **Breathlessness** | |  |
| --- | --- | --- |
| **Medcode** | **Readcode** | **Read term** |
| 101843 | 173a.00 | Borg Breathlessness Score: 10 maximal |
| 67566 | 173Y.00 | Borg Breathlessness Score: 9 very, very sev (almost maximal) |
| 108650 | 173g.00 | Breathlessness causing difficulty eating |
| 72334 | 173X.00 | Borg Breathlessness Score: 8 very severe (+) |
| 42287 | 173V.00 | Borg Breathlessness Score: 6 severe (+) |
| 70061 | 173W.00 | Borg Breathlessness Score: 7 very severe |
| 64049 | 173T.00 | Borg Breathlessness Score: 5 severe |
| 59860 | 173S.00 | Borg Breathlessness Score: 4 somewhat severe |
| 40813 | 173b.00 | Unable to complete a sentence in one breath |
| 107410 | 173f.00 | Anxiety about breathlessness |
| 57759 | 173Q.00 | Borg Breathlessness Score: 2 slight |
| 57193 | 173R.00 | Borg Breathlessness Score: 3 moderate |
| 22094 | 173F.00 | Short of breath dressing/undressing |
| 53771 | 173C.11 | Dyspnoea on exertion |
| 37704 | 2323 | O/E - orthopnoea |
| 9089 | 1735.11 | Orthopnoea symptom |
| 18116 | 173D.00 | Nocturnal dyspnoea |
| 24889 | 173G.00 | Breathless - strenuous exertion |
| 2563 | R060600 | [D]Respiratory distress |
| 21801 | 173Z.00 | Breathlessness NOS |
| 7683 | 1735 | Breathless - lying flat |
| 11451 | R060200 | [D]Orthopnoea |
| 31143 | 1734 | Breathless - at rest |
| 7000 | 2322 | O/E - dyspnoea |
| 6434 | 1736 | Paroxysmal nocturnal dyspnoea |
| 735 | R060D00 | [D]Breathlessness |
| 19429 | 173L.00 | MRC Breathlessness Scale: grade 5 |
| 12474 | 173C.12 | SOBOE |
| 7932 | 1733 | Breathless - mild exertion |
| 2931 | 1738 | Difficulty breathing |
| 5896 | 173..12 | Dyspnoea - symptom |
| 6326 | 1732 | Breathless - moderate exertion |
| 2575 | 173C.00 | Short of breath on exertion |
| 3092 | R060A00 | [D]Dyspnoea |
| 19430 | 173K.00 | MRC Breathlessness Scale: grade 4 |
| 5175 | 173..11 | Breathlessness symptom |
| 5349 | 173..13 | Shortness of breath symptom |
| 19426 | 173J.00 | MRC Breathlessness Scale: grade 3 |
| 1429 | 173..00 | Breathlessness |
| 741 | R060800 | [D]Shortness of breath |
| 19427 | 173I.00 | MRC Breathlessness Scale: grade 2 |
| 4822 | 1739 | Shortness of breath |

| **Peripheral oedema** | | |
| --- | --- | --- |
| **Medcode** | **Readcode** | **Read term** |
| 1284 | 22C3.00 | O/E - oedema of feet |
| 30309 | 183Z.00 | Oedema NOS |
| 12144 | 22CA.11 | O/E - arm oedema |
| 6651 | 22C..11 | O/E - swelling - oedema |
| 5919 | 1833.11 | Leg swelling symptom |
| 23570 | R023100 | [D]Anasarca |
| 61224 | 22C5.00 | O/E - oedema of thighs |
| 3158 | 183..00 | Oedema |
| 31747 | 22C6.00 | O/E - abdominal oedema |
| 9108 | 1837 | Pitting oedema |
| 7106 | 22C3.11 | O/E - foot oedema |
| 103579 | 183C.00 | Reduction in peripheral oedema |
| 11396 | R023400 | [D]Peripheral oedema |
| 19358 | 22C4.00 | O/E - oedema of legs |
| 10931 | 22C..00 | O/E - oedema |
| 14947 | R023300 | [D]Oedema, localized |
| 28419 | 22CZ.00 | O/E - oedema NOS |
| 29444 | 22CA.00 | O/E - oedema of arms |
| 98612 | 1839 | Oedema of calf |
| 20301 | R023000 | [D]Oedema, generalized |
| 14702 | R023z00 | [D]Oedema NOS |
| 6585 | 22C4.11 | O/E - leg oedema |
| 6047 | 183..11 | Oedema - symptom |
| 15044 | R023200 | [D]Dropsy |
| 4950 | R023.00 | [D]Oedema |
| 2140 | 183..12 | Swelling - oedema - symptom |
| 102720 | 183A.00 | Worsening peripheral oedema |
| 29264 | 22C4000 | Unilateral leg oedema |
| 19714 | 22C5.11 | O/E - thigh oedema |
| 1906 | 22C2.11 | O/E - ankle oedema |
| 7122 | 1832 | Ankle swelling |
| 20553 | 22C2.00 | O/E - oedema of ankles |
| 5889 | 1832.11 | Ankle swelling symptom |
| 43821 | ZR3R.00 | Cardiovasc Limits and Symptoms Profile ankle swelling score |
| 47344 | ZR3R.11 | CLASP ankle swelling score |
| 57963 | 388G.00 | Cardiovasc Limits and Symptoms Profile ankle swelling score |

| **Fatigue** |  |  |
| --- | --- | --- |
| **Medcode** | **Readcode** | **Read term** |
| 5794 | 168..00 | Tiredness symptom |
| 6242 | 168..11 | Fatigue - symptom |
| 5583 | 168..12 | Lethargy - symptom |
| 1816 | 168..13 | Malaise - symptom |
| 1404 | 1682 | Fatigue |
| 5751 | 1683 | Tired all the time |
| 15516 | 1683.11 | C/O - 'tired all the time' |
| 17736 | 1684 | Malaise/lethargy |
| 9823 | 1684.11 | C/O - debility - malaise |
| 9220 | 1688 | Exhaustion |
| 29292 | 168Z.00 | Tiredness symptom NOS |
| 7235 | E205.12 | Tired all the time |
| 9656 | Eu46011 | [X]Fatigue syndrome |
| 4546 | F286.00 | Chronic fatigue syndrome |
| 7529 | F286.11 | CFS - Chronic fatigue syndrome |
| 98512 | F286000 | Mild chronic fatigue syndrome |
| 97284 | F286100 | Moderate chronic fatigue syndrome |
| 98734 | F286200 | Severe chronic fatigue syndrome |
| 44215 | R007.00 | [D]Malaise and fatigue |
| 5658 | R007000 | [D]Malaise |
| 1688 | R007100 | [D]Fatigue |
| 5049 | R007200 | [D]Asthenia NOS |
| 9889 | R007211 | [D]General weakness |
| 1371 | R007300 | [D]Lethargy |
| 1147 | R007500 | [D]Tiredness |
| 23932 | R007z00 | [D]Malaise and fatigue NOS |
| 5814 | R007z11 | [D]Lassitude |

| **Abdominal bloating** | | |
| --- | --- | --- |
| **Medcode** | **Readcode** | **Read term** |
| 5150 | R073400 | [D]Bloating |
| 5821 | 19B..12 | Bloating symptom |
| 29567 | 19A2.00 | Abdomen feels bloated |

| **Pulmonary oedema** | | |
| --- | --- | --- |
| **Medcode** | **Readcode** | **Read term** |
| 102627 | 183B.00 | Worsening pulmonary oedema |
| 5155 | 23E1.00 | O/E - pulmonary oedema |
| 43618 | G581.12 | Pulmonary oedema - acute |
| 26082 | H541000 | Chronic pulmonary oedema |
| 7321 | H541z00 | Pulmonary oedema NOS |
| 558 | H584.00 | Acute pulmonary oedema unspecified |
| 5293 | H584z00 | Acute pulmonary oedema NOS |

| **Palpitations** | |  |
| --- | --- | --- |
| **Medcode** | **Readcode** | **Read term** |
| 20692 | 181..11 | Awareness of heartbeat |
| 10469 | 181..12 | Fluttering of heart |
| 326 | 181..00 | Palpitations |
| 29469 | 1813 | 'Bumping' of heart |
| 15616 | 181Z.00 | Palpitations NOS |
| 16170 | 1814 | 'Fluttering' of heart |
| 2975 | 1812 | Palpitations |

| **Chest pain** | |  |
| --- | --- | --- |
| **Medcode** | **Readcode** | **Read term** |
| 374 | 182..00 | Chest pain |
| 7346 | 1822 | Central chest pain |
| 1865 | 1823 | Precordial pain |
| 9698 | 1824 | Anterior chest wall pain |
| 1059 | 1825 | Pleuritic pain |
| 32612 | 1826 | Parasternal pain |
| 20490 | 1827 | Painful breathing -pleurodynia |
| 1283 | 1827.11 | Pleurodynia |
| 726 | 1828 | Atypical chest pain |
| 1228 | 1829 | Retrosternal pain |
| 18134 | 182A.00 | Chest pain on exertion |
| 2519 | 182B.00 | Rib pain |
| 8349 | 182B000 | Costal margin chest pain |
| 24704 | 182C.00 | Chest wall pain |
| 10370 | 182Z.00 | Chest pain NOS |
| 24321 | 1D22000 | Chest wall tenderness |
| 8264 | 8HTG.00 | Referred to acute chest pain clinic |
| 12509 | 8HTJ.00 | Referral to rapid access chest pain clinic |
| 9714 | 9N0f.00 | Seen in rapid access chest pain clinic |
| 32450 | G33z400 | Ischaemic chest pain |
| 2584 | R065.00 | [D]Chest pain |
| 544 | R065000 | [D]Chest pain, unspecified |
| 29490 | R065011 | [D] Retrosternal chest pain |
| 20481 | R065100 | [D]Precordial pain |
| 14823 | R065200 | [D]Anterior chest wall pain |
| 14819 | R065300 | [D]Painful respiration NOS |
| 18183 | R065400 | [D]Pleuritic pain |
| 24761 | R065500 | [D]Pleurodynia |
| 7878 | R065600 | [D]Chest discomfort |
| 15528 | R065700 | [D]Chest pressure |
| 1270 | R065800 | [D]Chest tightness |
| 19199 | R065900 | [D]Parasternal chest pain |
| 3518 | R065A00 | [D]Musculoskeletal chest pain |
| 9340 | R065B00 | [D]Non cardiac chest pain |
| 7844 | R065B14 | [D]Non-cardiac chest pain |
| 21082 | R065C00 | [D]Retrosternal chest pain |
| 50477 | R065D00 | [D]Central chest pain |
| 3796 | R065z00 | [D]Chest pain NOS |
| 53806 | Ryu0400 | [X]Other chest pain |

| **Pain** |  |  |
| --- | --- | --- |
| **Medcode** | **Readcode** | **Read term** |
| 3049 | J574700 | Anal pain |
| 93588 | 2C55.00 | O/E - painful splenomegaly |
| 526 | 1A53.11 | C/O - loin pain |
| 67672 | 2IA..00 | O/E - sign painful |
| 154 | N142.11 | Low back pain |
| 105257 | F369.00 | Complex regional pain syndrome |
| 103540 | R090P00 | [D]Functional abdominal pain syndrome |
| 109073 | 389Q.00 | Pain assessment tool completed |
| 286 | N094K12 | Hip pain |
| 4498 | R081000 | [D]Painful urination |
| 39146 | 1A58.11 | Female genital pain |
| 19020 | 25C..13 | O/E - lumbar pain on palpation |
| 19687 | 1M0..00 | Pain in upper limb |
| 36132 | 2H45.00 | O/E - joint movement painful |
| 22608 | 197C.00 | Lower abdominal pain |
| 21965 | ZLA2C00 | Seen by pain management nurse |
| 1666 | K583.13 | Period pains |
| 29400 | 1A5A.00 | C/O perineal pain |
| 50662 | Ryu1000 | [X]Pain localized to other parts of lower abdomen |
| 15039 | R041.00 | [D]Throat pain |
| 3978 | 197B.00 | Upper abdominal pain |
| 24832 | SP07300 | Internal prosthetic device causing pain |
| 2532 | N245400 | Calf pain |
| 24821 | 1974 | Right subcostal pain |
| 6629 | 29E..00 | O/E - pain sensation |
| 1181 | 1977 | Right iliac fossa pain |
| 1330 | N094512 | Hip joint pain |
| 110819 | 1DCF.00 | Gnawing pain |
| 3920 | R07A.00 | [D]Pain in oesophagus |
| 89917 | 7P21200 | Delivery of rehabilitation for pain syndromes |
| 42783 | 1L1..00 | Pain to call time |
| 109034 | 1M11000 | Ischaemic foot pain at rest |
| 5691 | 1963 | Non-colicky abdominal pain |
| 36040 | Eu45414 | [X]Somatoform pain disorder |
| 15435 | 1A5Z.00 | Genitourinary pain NOS |
| 3297 | SP2y200 | Postoperative pain |
| 42197 | Z1H2.00 | Pain rehabilitation |
| 2707 | 1A5C.00 | Pain in scrotum |
| 602 | K317000 | Mastodynia - pain in breast |
| 7294 | N240300 | Rheumatic pain |
| 111890 | 1M6..00 | Affective dimension of pain |
| 701 | 1975 | Left flank pain |
| 5673 | 1A58200 | Ovarian pain |
| 19360 | R090M00 | [D]Right lower quadrant pain |
| 36558 | 25C5.00 | O/E - abd. pain - R.lumbar |
| 51337 | 1964 | Shoulder pain from abdomen |
| 11609 | 1M1..00 | Pain in lower limb |
| 109179 | 1DCH.00 | Throbbing pain |
| 4544 | N245.00 | Pain in limb |
| 39867 | 29EZ.00 | O/E - pain sensation NOS |
| 6520 | N245000 | Hand pain |
| 9061 | R090L00 | [D]Left lower quadrant pain |
| 5899 | 1A53.12 | C/O - lumbar pain |
| 46856 | ZRb8.00 | Pain diary |
| 52971 | Ryu7000 | [X]Other chronic pain |
| 2025 | N245.14 | Hand pain |
| 100698 | 16ZB000 | Pain or discomfort |
| 1274 | N33A100 | Clavicle pain |
| 4617 | 1971 | Central abdominal pain |
| 93398 | 1M3..00 | Pain in face |
| 2600 | 1A53.13 | C/O - renal pain |
| 109209 | 1M50.00 | Constant pain |
| 36656 | Z1H1.12 | Controlling pain |
| 32539 | ZL17.00 | Under care of pain management specialist |
| 1335 | N142.00 | Pain in lumbar spine |
| 5796 | SP2y211 | Post-operative pain |
| 10084 | 1DC8.00 | Generalised pain [symptom] |
| 99477 | 1M01.00 | Pain in wrist |
| 7812 | 1962 | Colicky abdominal pain |
| 9517 | 1M10.00 | Knee pain |
| 15826 | F587100 | Otogenic pain |
| 46604 | ZL22C00 | Under care of pain management nurse |
| 43476 | ZLD2J00 | Discharge by pain management specialist |
| 5762 | N245300 | Pain in arm |
| 20357 | K197100 | Painful haematuria |
| 16546 | 2I18.11 | O/E - pain |
| 51210 | ZL62C00 | Referral to pain management nurse |
| 1946 | N245111 | Toe pain |
| 17799 | 1M11.00 | Foot pain |
| 855 | N245.13 | Foot pain |
| 8436 | R090H00 | [D]Upper abdominal pain |
| 94815 | 1ABD.00 | Painful erection |
| 24627 | 25C6.00 | O/E - abd. pain - umbilical |
| 29402 | N094411 | Hand joint pain |
| 11544 | N242300 | Neuropathic pain |
| 70001 | Z1G2111 | Managing pain |
| 6899 | 9N1k.00 | Seen in pain clinic |
| 7938 | K583.12 | Painful menstruation |
| 5780 | N245200 | Pain in leg |
| 2982 | 1978 | Left iliac fossa pain |
| 10902 | Z1H..00 | Pain management |
| 5806 | 2D7..00 | O/E - painful ear |
| 20359 | M2y1500 | Painful operation scar |
| 73315 | 2IA4.00 | O/E - sign very painful |
| 37101 | 25C4.00 | O/E - abd.pain-L.hypochondrium |
| 110285 | 1M60.00 | Punishing with pain |
| 29535 | L463500 | Pain on breast feeding |
| 111307 | 1M5..00 | Pattern of pain |
| 7726 | R090J00 | [D]Right upper quadrant pain |
| 17971 | SP07C00 | Pain due to shoulder joint prosthesis |
| 11906 | K273.11 | Erection - painful |
| 17324 | 1DC5.00 | Griping pain |
| 6831 | 8BAA.00 | Pain relief |
| 5840 | N143.11 | Acute back pain with sciatica |
| 2866 | J574F00 | Anorectal pain |
| 53766 | E278000 | Psychogenic pain unspecified |
| 24661 | 197A.00 | Generalised abdominal pain |
| 8600 | 1D18.00 | Pain from metastases |
| 11998 | N147211 | Pain in coccyx |
| 109208 | 1M51.00 | Intermittent pain |
| 15243 | R08z100 | [D]Vesical pain |
| 7490 | 197..11 | Flank pain |
| 109887 | 1M61.00 | Fearful with pain |
| 8931 | 1M...00 | Pain |
| 7300 | 1979 | Suprapubic pain |
| 17580 | 1DC6.00 | Tightening pain |
| 6644 | 1D13000 | C/O - pain in toes |
| 24796 | 16C4.00 | Back pain worse on sneezing |
| 17269 | 26C3.12 | Painful nipple |
| 18878 | 1944 | Painful swallowing |
| 103942 | F262C00 | Shortlast unilat neuralgifrm pain+conjunc inject+tearng synd |
| 109136 | 1DCJ.00 | Tender pain |
| 3258 | R040000 | [D]Facial pain |
| 5023 | N142.13 | Acute back pain - lumbar |
| 7056 | 1DC3.00 | Stabbing pain |
| 18374 | N239.11 | Myofascial pain syndrome |
| 5440 | N241012 | Muscle pain |
| 31062 | R090y00 | [D]Other specified abdominal pain |
| 34881 | Eu62y11 | [X]Chronic pain personality syndrome |
| 25200 | ZQ33.00 | Pain assessment |
| 177 | 1969 | Abdominal pain |
| 2522 | R00z200 | [D]Pain, generalized |
| 19322 | 1M13.00 | Ankle pain |
| 17079 | 16Z2.11 | Growing pains symptom |
| 6395 | 196..12 | Type of GIT pain - symptom |
| 96966 | N248000 | Myofascial pain syndrome |
| 1639 | K28z.11 | Pain in testis |
| 12189 | 16CA.00 | Mechanical low back pain |
| 109760 | 1M8..00 | Diabetic peripheral neuropathic pain |
| 26076 | 9Ok5.00 | Cancer pain and symptom management |
| 13018 | 196A.00 | Hunger pain |
| 29352 | 1969000 | Abdominal wall pain |
| 5960 | 197..13 | Site of abdominal pain |
| 48323 | Z1H1.13 | Giving pain relief |
| 14807 | R073.00 | [D]Flatulence, eructation and gas pain |
| 20475 | R090800 | [D]Suprapubic pain |
| 8541 | 19EC.00 | Painful defaecation |
| 4706 | N33A000 | Bony pelvic pain |
| 16868 | R090A00 | [D]Pain in left iliac fossa |
| 5787 | N245700 | Shoulder pain |
| 8362 | R090K00 | [D]Left upper quadrant pain |
| 105200 | F369.11 | Chronic regional pain syndrome |
| 5132 | 1A5B.00 | Pain in penis |
| 16078 | 1A57.00 | Pain in testicle |
| 38591 | Z23AN00 | Painful uterine contractions |
| 14852 | 1A58.00 | Pain in female genitalia |
| 10231 | 16C9.00 | Chronic low back pain |
| 11647 | 25C8.00 | O/E - abd. pain - R.iliac |
| 20640 | 25C..11 | O/E - epigastric pain on palp. |
| 37432 | 2D7Z.00 | O/E - painful ear NOS |
| 3338 | R090z00 | [D]Abdominal pain NOS |
| 19283 | R090N00 | [D]Nonspecific abdominal pain |
| 6112 | 1DC7.00 | Pricking pain |
| 42138 | 1M2..00 | Pain score |
| 14916 | 25CZ.00 | O/E -abd.pain on palpation NOS |
| 386 | 1CB3.00 | Throat pain |
| 50825 | 2D72.00 | O/E - ear auricle painful |
| 17602 | 26BD.00 | Intractable breast pain |
| 32957 | ZLEB.00 | Discharge from pain management service |
| 552 | N211z11 | Painful arc syndrome |
| 37565 | SyuKE11 | [X] Pain due to internal orthopaedic prosthesis |
| 4771 | R090600 | [D]Umbilical pain |
| 11979 | 1M00.11 | Elbow pain |
| 12774 | 1A5E.00 | Pain in vulva |
| 22049 | SP3y600 | Pain at injection site |
| 45921 | Z1H..11 | Managing pain |
| 7254 | 1DC1.00 | Burning pain |
| 2521 | N094311 | Wrist joint pain |
| 5923 | N145.11 | Acute back pain - unspecified |
| 96084 | N094F11 | Wrist pain |
| 19223 | 25C3.00 | O/E - abd. pain - epigastrium |
| 109741 | 1DCE.00 | Heavy pain |
| 17223 | 25C..12 | O/E - iliac pain on palpation |
| 7366 | 1CB3.11 | Pain in throat |
| 59352 | Z1H1.11 | Relieving pain |
| 554 | N094611 | Knee joint pain |
| 23872 | 1973 | Left subcostal pain |
| 30320 | Z1H1.00 | Pain control |
| 3191 | L16y500 | Abdominal pain in pregnancy |
| 9196 | 16Z2.00 | Growing pains |
| 3512 | N245.19 | Pain in buttock |
| 15180 | 25C..00 | O/E - abdo. pain on palpation |
| 9920 | R090G00 | [D]Pelvic and perineal pain |
| 8060 | 1DC2.00 | Aching pain |
| 5476 | N12..13 | Acute back pain - disc |
| 112826 | 1M62.00 | Sickening with pain |
| 16547 | K58y000 | Other pelvic pain - female |
| 1597 | J096.12 | Painful tongue |
| 73064 | 2IA2.00 | O/E - sign slightly painful |
| 44783 | 1D19.00 | Pain in lymph nodes after alcohol consumption |
| 54791 | N094D11 | Elbow joint pain |
| 11070 | 197A.11 | General abdominal pain-symptom |
| 52402 | Ryu1100 | [X]Other and unspecified abdominal pain |
| 14648 | 1A57.11 | Testicular pain |
| 95878 | N33C.00 | Complex regional pain syndrome type I |
| 105661 | 8CMW000 | Low back pain clinical pathway |
| 2494 | N245.15 | Heel pain |
| 4756 | 1A58000 | Vaginal pain |
| 4948 | N141.00 | Pain in thoracic spine |
| 7248 | R090G12 | [D] Perineal pain |
| 421 | 197..12 | Iliac fossa pain |
| 15213 | 2I18.00 | O/E - tenderness/pain |
| 4603 | SP07B00 | Pain due to knee joint prosthesis |
| 4627 | N245100 | Foot pain |
| 9811 | R090G11 | [D] Pelvic pain |
| 18016 | F336000 | Phantom limb syndrome with pain |
| 55007 | 1DC4.00 | Cutting pain |
| 4419 | N245500 | Axillary pain |
| 15288 | R090C00 | [D]Loin pain |
| 11718 | 196B.00 | Painful rectal bleeding |
| 9695 | 197D.00 | Right upper quadrant pain |
| 12156 | M2y1z12 | Scar painful |
| 107475 | 8HVk.00 | Private referral to pain management service |
| 1646 | N094211 | Elbow joint pain |
| 12160 | ZL98.00 | Seen by pain management specialist |
| 16328 | 22B6.00 | O/E - pain influenced posture |
| 4287 | K582.00 | Mittelschmerz - ovulation pain |
| 47797 | 29E2.00 | O/E - pain sensation reduced |
| 4733 | K28y800 | Pain in testis |
| 53478 | Z1H1200 | Minimising pain |
| 3000 | 1A58100 | Vulval pain |
| 5916 | N141.11 | Acute back pain - thoracic |
| 110070 | 1DCD.00 | Splitting pain |
| 123 | N131.00 | Cervicalgia - pain in neck |
| 41564 | R073z00 | [D]Flatulence, eructation and gas pain NOS |
| 1497 | R00z211 | [D]General aches and pains |
| 1339 | N245011 | Thumb pain |
| 16884 | N144011 | Thoracic nerve root pain |
| 31668 | 2IA..11 | O/E - painful sign |
| 18644 | 1DCA.00 | Rest pain |
| 97232 | Z1H6.00 | Pain control agreement |
| 109347 | 1M03.00 | Shoulder joint painful on external rotation |
| 1976 | 196..11 | Abdominal pain type |
| 8309 | N096.12 | Musculoskeletal pain - joints |
| 1286 | R040z11 | [D]Jaw pain |
| 48922 | Z1H3.00 | Pain management programme |
| 6704 | N145.12 | Back pain, unspecified |
| 93531 | F347.00 | Complex regional pain syndrome type II |
| 37634 | Z714200 | Back pain prevention training |
| 61911 | 9b8F.00 | Pain management (specialty) |
| 110496 | 1M52.00 | Chronic pain |
| 3324 | 16C6.00 | Back pain without radiation NOS |
| 2048 | 1D13.11 | Pain |
| 2767 | J574800 | Rectal pain |
| 100639 | 1M4..00 | Central post-stroke pain |
| 11017 | 8BAO.00 | Pain and symptom management |
| 107428 | 9NNh.00 | Under care of pain management specialist |
| 5783 | 1B84.00 | Has eye pain |
| 1219 | N245.16 | Leg pain |
| 42211 | 25C9.00 | O/E - abd. pain - hypogastrium |
| 1457 | J046400 | Temporomandibular joint-pain-dysfunction syndrome |
| 67678 | 2IAZ.00 | O/E - sign painful NOS |
| 14886 | N094711 | Ankle joint pain |
| 15234 | 1DC..00 | Pain character |
| 108998 | 1DCG.00 | Cramping pain |
| 109217 | 1M11100 | Ischaemic foot pain when walking |
| 917 | N33A.00 | Bone pain |
| 11698 | 196C.00 | Painless rectal bleeding |
| 106388 | 66n..00 | Chronic pain review |
| 6928 | 2H23.11 | O/E - painful arc |
| 1336 | R090B00 | [D]Groin pain |
| 6357 | 197..14 | Subcostal pain |
| 13670 | ZL58.00 | Referral to pain management specialist |
| 6313 | 1D13111 | C/O - pain in big toe |
| 8599 | 1DC9.00 | Shooting pain |
| 2781 | 1A59.00 | C/O pelvic pain |
| 8433 | 1A5D.00 | Urethral pain |
| 14989 | 196..00 | Type of GIT pain |
| 5920 | 1B84.11 | Pain in eye |
| 9728 | 1DCZ.00 | Pain character NOS |
| 6424 | 1A5..00 | Genitourinary pain |
| 28285 | R073200 | [D]Gas pain (abdominal) |
| 6166 | N094W00 | Anterior knee pain |
| 92389 | Z7CM300 | Agnosia for pain |
| 110568 | Z1H7.00 | Partnership in pain control |
| 628 | R090700 | [D]Hypochondrial pain |
| 33549 | F587200 | Referred ear pain |
| 198 | N245.17 | Shoulder pain |
| 6747 | F302.00 | Atypical face pain |
| 9105 | 1D13100 | C/O - pain in hallux |
| 70440 | 8B3g.00 | Pain to thrombolysis time |
| 29922 | 197Z.00 | Site of GIT pain NOS |
| 11040 | SP07A00 | Pain due to hip joint prosthesis |
| 3763 | 16C5.00 | C/O - low back pain |
| 3322 | N245012 | Finger pain |
| 35785 | F372100 | Chronic painful diabetic neuropathy |
| 52859 | Ryu4200 | [X]Painful micturition, unspecified |
| 17112 | N134.13 | Cervical root pain |
| 24060 | R00zC00 | [D]Chronic intractable pain |
| 44484 | 196Z.00 | Type of GIT pain NOS |
| 48078 | F372000 | Acute painful diabetic neuropathy |
| 3086 | 1976 | Right flank pain |
| 46297 | Z246311 | Onset of labour pains |
| 7014 | 1A54.12 | C/O - ureteric pain |
| 5754 | 1BA5.11 | Pain in sinuses |
| 5908 | 2252 | O/E - in pain |
| 30179 | Eu45400 | [X]Persistent somatoform pain disorder |
| 61479 | 2IA3.00 | O/E - sign moderately painful |
| 1866 | N245.18 | Thigh pain |
| 25118 | 197..00 | Site of GIT pain |
| 21618 | R079.00 | [D] Defaecation painful |
| 1455 | R01z200 | [D]Musculoskeletal pain |
| 2792 | M2y1z11 | Painful scar |
| 3266 | K583.11 | Painful menorrhoea |
| 5864 | N094.00 | Pain in joint - arthralgia |
| 40556 | 29E1.00 | O/E - pain sensation normal |
| 5056 | R040z00 | [D]Pain in head NOS |
| 2234 | R090E00 | [D]Recurrent acute abdominal pain |
| 2397 | N094111 | Shoulder joint pain |
| 59367 | Z1H1100 | Preventing pain |
| 103617 | 3880000 | Visual analogue pain scale |
| 2545 | 8BAB.00 | Pain control |
| 12639 | 25C2.00 | O/E - abd.pain-R.hypochondrium |
| 822 | N245.12 | Arm pain |
| 5966 | K28y811 | Testicular pain |
| 2098 | F4Kz100 | Eye pain NOS |
| 9651 | K197000 | Painless haematuria |
| 542 | R090500 | [D]Epigastric pain |
| 1258 | N245.11 | Ankle pain |
| 108855 | 1M02.00 | Shoulder joint painful on movement |
| 290 | 1972 | Epigastric pain |
| 25630 | 25C7.00 | O/E - abd. pain - L.lumbar |
| 5880 | 1D13.00 | C/O: a pain |
| 42040 | ZRrA111 | VAPS - Visual analogue pain scale |
| 16806 | R090900 | [D]Pain in right iliac fossa |
| 9682 | 8HTH.00 | Referral to back pain clinic |
| 10389 | 1M12.00 | Anterior knee pain |
| 21583 | 25CA.00 | O/E - abd. pain - L.iliac |
| 1135 | F587.11 | Ear pain |
| 8029 | R00zB00 | [D]Acute pain |
| 11962 | 1M00.00 | Pain in elbow |
| 15818 | 2D73.00 | O/E - pain over mastoid |
| 35744 | N131.11 | Pain in cervical spine |
| 1936 | 8H69.00 | Refer to pain clinic |
| 1763 | R090.00 | [D]Abdominal pain |

| **Depression** | |  |
| --- | --- | --- |
| **Medcode** | **Readcode** | **Read term** |
| 9796 | 1B1U.00 | Symptoms of depression |
| 10438 | 1B1U.11 | Depressive symptoms |
| 10015 | 1BT..00 | Depressed mood |
| 8928 | 1BT..11 | Low mood |
| 26028 | 1BT..12 | Sad mood |
| 100977 | 1JJ..00 | Suspected depression |
| 44848 | 8BK0.00 | Depression management programme |
| 30483 | 8CAa.00 | Patient given advice about management of depression |
| 32841 | 8HHq.00 | Referral for guided self-help for depression |
| 112682 | 8IH5200 | Referral for guided self-help for depression declined |
| 27677 | E001300 | Presenile dementia with depression |
| 44674 | E002.00 | Senile dementia with depressive or paranoid features |
| 21887 | E002100 | Senile dementia with depression |
| 41089 | E002z00 | Senile dementia with depressive or paranoid features NOS |
| 43292 | E004300 | Arteriosclerotic dementia with depression |
| 46244 | E02y300 | Drug-induced depressive state |
| 2560 | E11..12 | Depressive psychoses |
| 10610 | E112.00 | Single major depressive episode |
| 5879 | E112.11 | Agitated depression |
| 6546 | E112.12 | Endogenous depression first episode |
| 6950 | E112.13 | Endogenous depression first episode |
| 595 | E112.14 | Endogenous depression |
| 34390 | E112000 | Single major depressive episode, unspecified |
| 16506 | E112100 | Single major depressive episode, mild |
| 15155 | E112200 | Single major depressive episode, moderate |
| 15219 | E112300 | Single major depressive episode, severe, without psychosis |
| 32159 | E112400 | Single major depressive episode, severe, with psychosis |
| 43324 | E112500 | Single major depressive episode, partial or unspec remission |
| 7011 | E112z00 | Single major depressive episode NOS |
| 15099 | E113.00 | Recurrent major depressive episode |
| 6932 | E113.11 | Endogenous depression - recurrent |
| 35671 | E113000 | Recurrent major depressive episodes, unspecified |
| 29342 | E113100 | Recurrent major depressive episodes, mild |
| 14709 | E113200 | Recurrent major depressive episodes, moderate |
| 25697 | E113300 | Recurrent major depressive episodes, severe, no psychosis |
| 24171 | E113400 | Recurrent major depressive episodes, severe, with psychosis |
| 56273 | E113500 | Recurrent major depressive episodes,partial/unspec remission |
| 6482 | E113700 | Recurrent depression |
| 25563 | E113z00 | Recurrent major depressive episode NOS |
| 17385 | E114.11 | Manic-depressive - now manic |
| 12831 | E115.11 | Manic-depressive - now depressed |
| 60178 | E11y.00 | Other and unspecified manic-depressive psychoses |
| 11596 | E11y000 | Unspecified manic-depressive psychoses |
| 27491 | E11y200 | Atypical depressive disorder |
| 70399 | E11y300 | Other mixed manic-depressive psychoses |
| 33426 | E11yz00 | Other and unspecified manic-depressive psychoses NOS |
| 9183 | E11z200 | Masked depression |
| 8478 | E130.00 | Reactive depressive psychosis |
| 17770 | E130.11 | Psychotic reactive depression |
| 1055 | E135.00 | Agitated depression |
| 1131 | E204.00 | Neurotic depression reactive type |
| 1533 | E290.00 | Brief depressive reaction |
| 36246 | E290z00 | Brief depressive reaction NOS |
| 16632 | E291.00 | Prolonged depressive reaction |
| 324 | E2B..00 | Depressive disorder NEC |
| 2972 | E2B0.00 | Postviral depression |
| 4323 | E2B1.00 | Chronic depression |
| 20785 | Eu20400 | [X]Post-schizophrenic depression |
| 11055 | Eu25100 | [X]Schizoaffective disorder, depressive type |
| 41022 | Eu25112 | [X]Schizophreniform psychosis, depressive type |
| 1531 | Eu31.11 | [X]Manic-depressive illness |
| 6710 | Eu31.12 | [X]Manic-depressive psychosis |
| 66153 | Eu31.13 | [X]Manic-depressive reaction |
| 4639 | Eu32.00 | [X]Depressive episode |
| 9055 | Eu32.11 | [X]Single episode of depressive reaction |
| 18510 | Eu32.12 | [X]Single episode of psychogenic depression |
| 7604 | Eu32.13 | [X]Single episode of reactive depression |
| 11717 | Eu32000 | [X]Mild depressive episode |
| 9211 | Eu32100 | [X]Moderate depressive episode |
| 22806 | Eu32212 | [X]Single episode major depression w'out psychotic symptoms |
| 59386 | Eu32213 | [X]Single episode vital depression w'out psychotic symptoms |
| 12099 | Eu32300 | [X]Severe depressive episode with psychotic symptoms |
| 24117 | Eu32311 | [X]Single episode of major depression and psychotic symptoms |
| 52678 | Eu32312 | [X]Single episode of psychogenic depressive psychosis |
| 24112 | Eu32313 | [X]Single episode of psychotic depression |
| 28863 | Eu32314 | [X]Single episode of reactive depressive psychosis |
| 10667 | Eu32400 | [X]Mild depression |
| 98346 | Eu32500 | [X]Major depression, mild |
| 98252 | Eu32600 | [X]Major depression, moderately severe |
| 98414 | Eu32700 | [X]Major depression, severe without psychotic symptoms |
| 98417 | Eu32800 | [X]Major depression, severe with psychotic symptoms |
| 6854 | Eu32y00 | [X]Other depressive episodes |
| 10720 | Eu32y11 | [X]Atypical depression |
| 56609 | Eu32y12 | [X]Single episode of masked depression NOS |
| 2970 | Eu32z00 | [X]Depressive episode, unspecified |
| 543 | Eu32z11 | [X]Depression NOS |
| 3291 | Eu32z12 | [X]Depressive disorder NOS |
| 28248 | Eu32z13 | [X]Prolonged single episode of reactive depression |
| 5987 | Eu32z14 | [X] Reactive depression NOS |
| 3292 | Eu33.00 | [X]Recurrent depressive disorder |
| 8851 | Eu33.11 | [X]Recurrent episodes of depressive reaction |
| 19696 | Eu33.12 | [X]Recurrent episodes of psychogenic depression |
| 8902 | Eu33.13 | [X]Recurrent episodes of reactive depression |
| 28756 | Eu33.14 | [X]Seasonal depressive disorder |
| 29784 | Eu33000 | [X]Recurrent depressive disorder, current episode mild |
| 29520 | Eu33100 | [X]Recurrent depressive disorder, current episode moderate |
| 11329 | Eu33211 | [X]Endogenous depression without psychotic symptoms |
| 11252 | Eu33212 | [X]Major depression, recurrent without psychotic symptoms |
| 73991 | Eu33214 | [X]Vital depression, recurrent without psychotic symptoms |
| 23731 | Eu33311 | [X]Endogenous depression with psychotic symptoms |
| 32941 | Eu33313 | [X]Recurr severe episodes/major depression+psychotic symptom |
| 31757 | Eu33314 | [X]Recurr severe episodes/psychogenic depressive psychosis |
| 16861 | Eu33315 | [X]Recurrent severe episodes of psychotic depression |
| 37764 | Eu33316 | [X]Recurrent severe episodes/reactive depressive psychosis |
| 47731 | Eu33y00 | [X]Other recurrent depressive disorders |
| 44300 | Eu33z00 | [X]Recurrent depressive disorder, unspecified |
| 8584 | Eu34111 | [X]Depressive neurosis |
| 10290 | Eu34112 | [X]Depressive personality disorder |
| 7737 | Eu34113 | [X]Neurotic depression |
| 19054 | Eu3y111 | [X]Recurrent brief depressive episodes |
| 29527 | R007z13 | [D]Postoperative depression |

| **Anxiety** |  |  |
| --- | --- | --- |
| **Medcode** | **Readcode** | **Read term** |
| 23598 | E201300 | Hysterical tremor |
| 15292 | E262000 | Cardiac neurosis |
| 15224 | E263z00 | Psychogenic skin symptoms NOS |
| 57877 | Eu45100 | [X]Undifferentiated somatoform disorder |
| 31672 | E202900 | Fear of crowds |
| 966 | E207.00 | Hypochondriasis |
| 16415 | E293.00 | Adjustment reaction with predominant disturbance of conduct |
| 30961 | E262300 | Psychogenic cardiovascular disorder |
| 10390 | E202D00 | Fear of death |
| 191 | E278100 | Tension headache |
| 20634 | Eu42000 | [X]Predominantly obsessional thoughts or ruminations |
| 22721 | Eu42z00 | [X]Obsessive-compulsive disorder, unspecified |
| 14780 | E20z.00 | Neurotic disorder NOS |
| 26138 | E28z.00 | Acute stress reaction NOS |
| 34696 | E201400 | Hysterical paralysis |
| 15284 | E262200 | Neurocirculatory asthenia |
| 35914 | E293100 | Adjustment reaction with antisocial behaviour |
| 31515 | Eu43z00 | [X]Reaction to severe stress, unspecified |
| 62002 | Eu45y00 | [X]Other somatoform disorders |
| 7537 | Eu45200 | [X]Hypochondriacal disorder |
| 104891 | E293z00 | Adjustment reaction with predominant disturbance conduct NOS |
| 28090 | Eu46.00 | [X]Other neurotic disorders |
| 43050 | E20yz00 | Other neurotic disorder NOS |
| 89237 | E267.00 | Psychogenic symptom of special sense organ |
| 66398 | E293200 | Adjustment reaction with destructiveness |
| 68379 | E265.00 | Psychogenic genitourinary tract symptoms |
| 27390 | E29y400 | Adjustment reaction due to hospitalisation |
| 64166 | Eu44y00 | [X]Other dissociative [conversion] disorders |
| 73547 | E265z00 | Psychogenic genitourinary tract symptom NOS |
| 23354 | E201z00 | Hysteria NOS |
| 5067 | E26z.00 | Psychosomatic disorder NOS |
| 56141 | Eu44200 | [X]Dissociative stupor |
| 15551 | E282.00 | Acute stupor state due to acute stress reaction |
| 24847 | E283100 | Acute posttrauma stress state |
| 15371 | E264300 | Psychogenic diarrhoea |
| 39747 | Eu44300 | [X]Trance and possession disorders |
| 15665 | E292z00 | Adjustment reaction with disturbance of other emotion NOS |
| 49628 | Eu46z00 | [X]Neurotic disorder, unspecified |
| 48906 | Eu44z00 | [X]Dissociative [conversion] disorder, unspecified |
| 32034 | E261000 | Psychogenic air hunger |
| 12228 | E211100 | Hypomanic personality disorder |
| 11607 | Eu43000 | [X]Acute stress reaction |
| 24212 | E292.00 | Adjustment reaction, predominant disturbance other emotions |
| 56800 | E260100 | Psychogenic torticollis |
| 3208 | E203.00 | Obsessive-compulsive disorders |
| 12838 | E202200 | Agoraphobia without mention of panic attacks |
| 29707 | E283z00 | Other acute stress reaction NOS |
| 2188 | E201.00 | Hysteria |
| 2366 | E202C00 | Dental phobia |
| 53362 | E29y000 | Concentration camp syndrome |
| 35632 | E29y200 | Adjustment reaction with physical symptoms |
| 71437 | E264z00 | Psychogenic gastrointestinal tract symptom NOS |
| 18049 | Eu45.00 | [X]Somatoform disorders |
| 40994 | Eu44000 | [X]Dissociative amnesia |
| 45603 | E294.00 | Adjustment reaction with disturbance emotion and conduct |
| 44739 | E201200 | Hysterical deafness |
| 55781 | E265300 | Psychogenic dysuria |
| 39518 | E20y200 | Other occupational neurosis |
| 276 | E28..00 | Acute reaction to stress |
| 62400 | E26y.00 | Other psychogenic malfunction |
| 37669 | E29z.00 | Adjustment reaction NOS |
| 23327 | E292100 | Adolescent emancipation disorder |
| 96391 | E26yz00 | Other psychogenic malfunction NOS |
| 15321 | E20y000 | Somatization disorder |
| 32387 | E29y100 | Other post-traumatic stress disorder |
| 20245 | E283000 | Acute situational disturbance |
| 4775 | E201800 | Hysterical fugue |
| 20053 | E261300 | Psychogenic hyperventilation |
| 38640 | E283.00 | Other acute stress reactions |
| 1907 | E202.00 | Phobic disorders |
| 4963 | E264000 | Psychogenic aerophagy |
| 39826 | Eu44100 | [X]Dissociative fugue |
| 41455 | E29y.00 | Other adjustment reactions |
| 3438 | E201100 | Hysterical blindness |
| 30179 | Eu45400 | [X]Persistent somatoform pain disorder |
| 22019 | Eu42100 | [X]Predominantly compulsive acts [obsessional rituals] |
| 40311 | E278.00 | Psychalgia |
| 44547 | E265200 | Psychogenic dysmenorrhea |
| 41572 | E201000 | Hysteria unspecified |
| 56966 | Eu44500 | [X]Dissociative convulsions |
| 4167 | E202A00 | Fear of flying |
| 3685 | E20y100 | Writer's cramp neurosis |
| 29461 | E263.00 | Psychogenic skin symptoms |
| 19921 | E29y500 | Other adjustment reaction with withdrawal |
| 23490 | E201A00 | Dissociative reaction unspecified |
| 4269 | E201700 | Hysterical amnesia |
| 14729 | E202z00 | Phobic disorder NOS |
| 15939 | E264500 | Psychogenic constipation |
| 11336 | Eu43200 | [X]Adjustment disorders |
| 2030 | E203100 | Obsessional neurosis |
| 12508 | Eu40300 | [X]Needle phobia |
| 5652 | E210.00 | Paranoid personality disorder |
| 5678 | E203000 | Compulsive neurosis |
| 1510 | E202B00 | Cancer phobia |
| 72171 | E20y300 | Psychasthenic neurosis |
| 20109 | E265100 | Psychogenic vaginismus |
| 7716 | E29y300 | Elective mutism due to an adjustment reaction |
| 54373 | E278z00 | Psychalgia NOS |
| 48561 | E260000 | Psychogenic paralysis |
| 9686 | E2...00 | Neurotic, personality and other nonpsychotic disorders |
| 11098 | Eu43.00 | [X]Reaction to severe stress, and adjustment disorders |
| 23808 | Eu4..00 | [X]Neurotic, stress - related and somoform disorders |
| 21753 | Eu43y00 | [X]Other reactions to severe stress |
| 5249 | E20..00 | Neurotic disorders |
| 31422 | E264.00 | Psychogenic gastrointestinal tract symptoms |
| 58013 | E292500 | Culture shock |
| 41615 | E261500 | Psychogenic aphonia |
| 5304 | Eu42.00 | [X]Obsessive - compulsive disorder |
| 38809 | Eu42y00 | [X]Other obsessive-compulsive disorders |
| 51497 | E211z00 | Affective personality disorder NOS |
| 2826 | E29..00 | Adjustment reaction |
| 5305 | E206.00 | Depersonalisation syndrome |
| 3361 | E205.00 | Neurasthenia - nervous debility |
| 14979 | E211.00 | Affective personality disorder |
| 27633 | Eu44600 | [X]Dissociative anaesthesia and sensory loss |
| 16178 | E211000 | Unspecified affective personality disorder |
| 9785 | Eu40200 | [X]Specific (isolated) phobias |
| 31957 | E202400 | Social phobia, fear of public speaking |
| 2775 | E290000 | Grief reaction |
| 18399 | Eu42200 | [X]Mixed obsessional thoughts and acts |
| 2871 | E264200 | Cyclical vomiting - psychogenic |
| 15483 | E261100 | Psychogenic cough |
| 23869 | E284.00 | Stress reaction causing mixed disturbance of emotion/conduct |
| 15566 | E203z00 | Obsessive-compulsive disorder NOS |
| 34664 | E261z00 | Psychogenic respiratory symptom NOS |
| 15959 | E263000 | Psychogenic pruritus |
| 42000 | E20y.00 | Other neurotic disorders |
| 38134 | E261.00 | Psychogenic respiratory symptoms |
| 3869 | E264400 | Psychogenic dyspepsia |
| 41038 | Eu45300 | [X]Somatoform autonomic dysfunction |
| 15034 | E262z00 | Psychogenic cardiovascular symptom NOS |
| 53766 | E278000 | Psychogenic pain unspecified |
| 12453 | Eu45500 | [X]Globus pharyngeus |
| 16484 | E201500 | Hysterical seizures |
| 44212 | E260.00 | Psychogenic musculoskeletal symptoms |
| 24439 | Eu45000 | [X]Somatization disorder |
| 16199 | E202300 | Social phobia, fear of eating in public |
| 22136 | Eu44.00 | [X]Dissociative [conversion] disorders |
| 42737 | E281.00 | Acute fugue state due to acute stress reaction |
| 16561 | Eu46000 | [X]Neurasthenia |
| 23413 | E261400 | Psychogenic yawning |
| 34978 | Eu44400 | [X]Dissociative motor disorders |
| 4171 | Eu43100 | [X]Post - traumatic stress disorder |
| 45205 | E278200 | Psychogenic backache |
| 88758 | Eu44700 | [X]Mixed dissociative [conversion] disorders |
| 15035 | E260z00 | Psychogenic musculoskeletal symptoms NOS |
| 29322 | E201B00 | Compensation neurosis |
| 29448 | E262.00 | Psychogenic cardiovascular symptoms |
| 9265 | Eu46100 | [X]Depersonalization - derealization syndrome |
| 47809 | E261200 | Psychogenic hiccough |
| 15431 | E201600 | Other conversion disorder |
| 28106 | E202600 | Acrophobia |
| 28938 | E202700 | Animal phobia |
| 6075 | E293000 | Adjustment reaction with aggression |
| 23462 | E29yz00 | Other adjustment reactions NOS |
| 9125 | 8G94.00 | Anxiety management training |
| 4199 | E26..00 | Physiological malfunction arising from mental factors |
| 99609 | ZS7C700 | Post-traumatic mutism |
| 48671 | Eu45z00 | [X]Somatoform disorder, unspecified |
| 48588 | E292y00 | Adjustment reaction with mixed disturbance of emotion |
| 12635 | Eu40214 | [X]Simple phobia |
| 1582 | E205.11 | Nervous exhaustion |
| 9944 | E202.12 | Phobic anxiety |
| 10723 | R2y2.12 | [D]Nervous tension |
| 28408 | 2J4..00 | Worried well |
| 50191 | Eu41113 | [X]Anxiety state |
| 7999 | Z4L1.00 | Anxiety counselling |
| 62935 | Z4I7100 | Recognising anxiety |
| 93401 | 1B13.12 | Anxious |
| 5902 | 1B13.11 | Anxiousness - symptom |
| 20375 | ZV65511 | [V]'Worried well' |
| 11890 | 1B1V.00 | C/O - panic attack |
| 19000 | 225J.00 | O/E - panic attack |
| 6408 | Eu41011 | [X]Panic attack |
| 7222 | Eu40z11 | [X]Phobia NOS |
| 2524 | 1BK..00 | Worried |
| 29608 | 1B12.00 | 'Nerves' - nervousness |
| 28167 | Eu41y11 | [X]Anxiety hysteria |
| 21431 | Eu46z11 | [X]Neurosis NOS |
| 791 | E20z.11 | Nervous breakdown |
| 26295 | Z4I7211 | Reducing anxiety |
| 8424 | Eu60600 | [X]Anxious [avoidant] personality disorder |
| 8725 | 2259 | O/E - nervous |
| 462 | E200111 | Panic attack |
| 11280 | Eu40213 | [X]Claustrophobia |
| 4081 | Eu41012 | [X]Panic state |
| 131 | 1B13.00 | Anxiousness |
| 14890 | Eu40012 | [X]Panic disorder with agoraphobia |
| 20773 | Eu05400 | [X]Organic anxiety disorder |
| 514 | 1B12.12 | Tension - nervous |
| 962 | Eu41111 | [X]Anxiety neurosis |
| 101422 | 16ZB100 | Feeling low or worried |
| 28381 | Z4I7200 | Alleviating anxiety |
| 2509 | R2y2.00 | [D]Nervousness |
| 35825 | Eu41112 | [X]Anxiety reaction |
| 13124 | 2258 | O/E - anxious |
| 107410 | 173f.00 | Anxiety about breathlessness |
| 22159 | Z4I7.00 | Acknowledging anxiety |
| 25638 | Eu41z11 | [X]Anxiety NOS |
| 17687 | Eu51511 | [X]Dream anxiety disorder |
| 6221 | E292000 | Separation anxiety disorder |
| 4069 | E200100 | Panic disorder |
| 23838 | Eu41z00 | [X]Anxiety disorder, unspecified |
| 11940 | E280.00 | Acute panic state due to acute stress reaction |
| 44321 | Eu41300 | [X]Other mixed anxiety disorders |
| 1758 | E200400 | Chronic anxiety |
| 6939 | E200000 | Anxiety state unspecified |
| 4659 | E200200 | Generalised anxiety disorder |
| 4634 | E200500 | Recurrent anxiety |
| 4534 | E200z00 | Anxiety state NOS |
| 2571 | Eu40000 | [X]Agoraphobia |
| 11602 | Eu40100 | [X]Social phobias |
| 18603 | E202500 | Social phobia, fear of public washing |
| 5385 | Eu41.00 | [X]Other anxiety disorders |
| 1723 | E202800 | Claustrophobia |
| 56924 | E292400 | Adjustment reaction with anxious mood |
| 2300 | E202000 | Phobia unspecified |
| 8205 | Eu41000 | [X]Panic disorder [episodic paroxysmal anxiety] |
| 10344 | Eu41100 | [X]Generalized anxiety disorder |
| 27685 | Eu40y00 | [X]Other phobic anxiety disorders |
| 9386 | Eu40.00 | [X]Phobic anxiety disorders |
| 24066 | Eu41y00 | [X]Other specified anxiety disorders |
| 636 | E200.00 | Anxiety states |
| 34064 | Eu40z00 | [X]Phobic anxiety disorder, unspecified |
| 3076 | E202100 | Agoraphobia with panic attacks |

### **S3 Measurement of exposures (symptoms) and time-horizons for each landmark time**

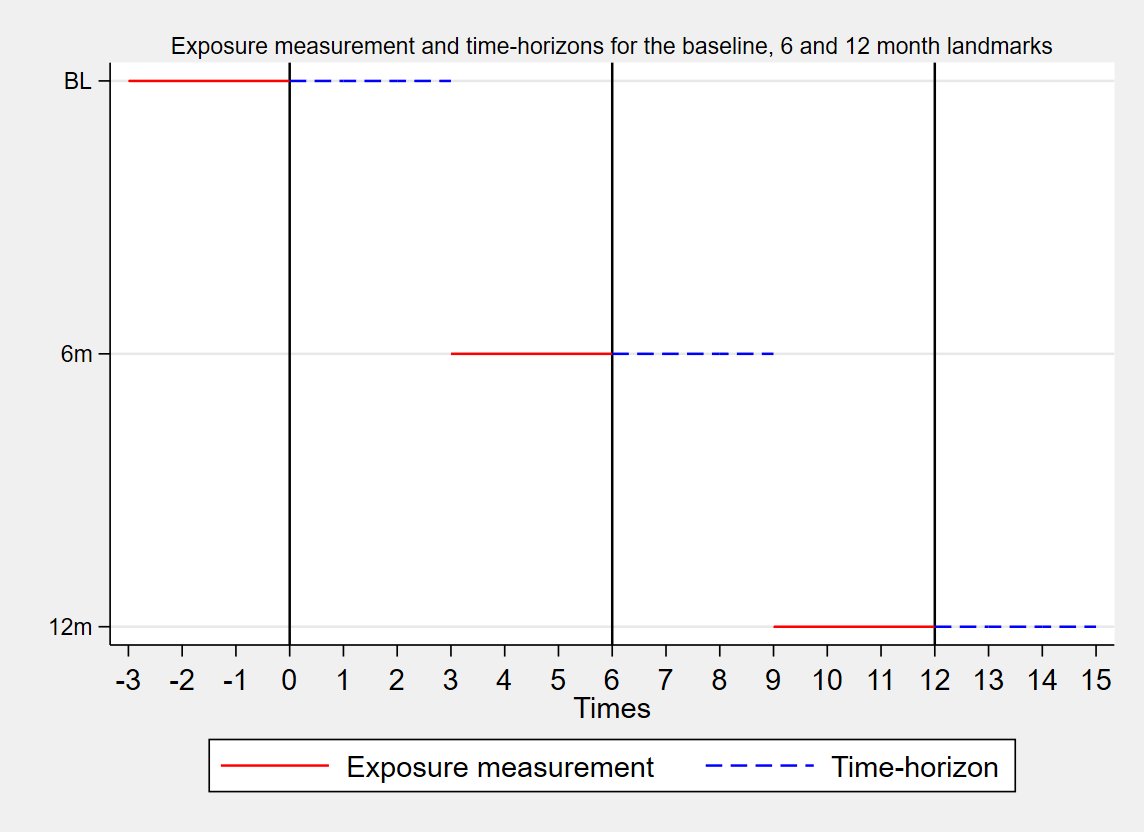

**S3 Table: Unadjusted and adjusted effect estimates of symptoms and outcomes (multiple imputed data)**

|  |  | **Baseline** | | **6 months** | | **12 months** | |
| --- | --- | --- | --- | --- | --- | --- | --- |
|  |  | **HR (95% CI)** | | **HR (95% CI)** | | **HR (95% CI)** | |
| **Outcome** | **Symptom** | **Unadjusted** | **Adjusted** | **Unadjusted** | **Adjusted** | **Unadjusted** | **Adjusted** |
| All-cause admission | SOB | 1.03 (1.01, 1.04) | 1.00 (0.97, 1.04) | 1.78 (1.67, 1.89) | **1.29 (1.21, 1.37)** | 1.84 (1.71, 1.98) | **1.30 (1.21, 1.40)** |
|  | Oedema | 1.10 (1.08, 1.13) | 1.04 (1.00, 1.09) | 1.49 (1.37, 1.63) | **1.19 (1.09, 1.30)** | 1.63 (1.47, 1.81) | **1.27 (1.15, 1.41)** |
|  | Ankle swelling | 1.04 (1.00, 1.07) | 1.02 (0.94, 1.10) | 1.33 (1.12, 1.60) | 1.12 (0.93, 1.34) | 1.68 (1.38, 2.04) | **1.47 (1.21, 1.79)** |
|  | Fatigue | 1.18 (1.14, 1.23) | **1.11 (1.02, 1.20)** | 1.49 (1.30, 1.72) | **1.24 (1.07, 1.43)** | 1.81 (1.55, 2.11) | **1.45 (1.24, 1.69)** |
|  | Chest pain | 0.91 (0.85, 0.97) | **1.11 (1.05, 1.17)** | 1.83 (1.66, 2.02) | **1.13 (1.02, 1.25)** | 1.88 (1.68, 2.11) | **1.14 (1.02, 1.28)** |
|  | Anxiety | 1.07 (1.02, 1.12) | 1.02 (0.92, 1.14) | 1.47 (1.22, 1.76) | **1.25 (1.04, 1.50)** | 1.74 (1.44, 2.10) | **1.28 (1.06, 1.55)** |
|  | Depression | 1.44 (1.38, 1.49) | **1.26 (1.15, 1.39)** | 1.68 (1.44, 1.96) | **1.46 (1.25, 1.70)** | 1.50 (1.25, 1.81) | **1.33 (1.10, 1.61)** |
|  | Pain | 1.16 (1.14, 1.18) | **1.09 (1.05, 1.14)** | 1.32 (1.24, 1.40) | **1.11 (1.05, 1.18)** | 1.36 (1.27, 1.45) | **1.16 (1.08, 1.24)** |
| HF admission | SOB | 1.11 (1.09, 1.14) | **1.18 (1.12, 1.26)** | 2.45 (2.14, 2.80) | **1.72 (1.50, 1.98)** | 2.97 (2.53, 3.49) | **1.99 (1.68, 2.35)** |
|  | Oedema | 1.33 (1.30, 1.37) | **1.14 (1.05, 1.24)** | 1.97 (1.62, 2.39) | **1.43 (1.18, 1.74)** | 2.36 (1.87, 2.99) | **1.64 (1.29, 2.08)** |
|  | Ankle swelling | 1.18 (1.12, 1.24) | 1.10 (0.95, 1.27) | 1.73 (1.16, 2.57) | 1.39 (0.93, 2.07) | 1.67 (0.98, 2.82) | 1.37 (0.81, 2.32) |
|  | Fatigue | 1.27 (1.21, 1.34) | 1.16 (0.99, 1.35) | 1.79 (1.30, 2.47) | **1.41 (1.02, 1.95)** | 2.36 (1.65, 3.37) | **1.78 (1.24, 2.56)** |
|  | Chest pain | 0.81 (0.73, 0.90) | 1.00 (0.90, 1.11) | 1.57 (1.21, 2.03) | 0.92 (0.70, 1.20) | 1.76 (1.29, 2.39) | 1.02 (0.75, 1.41) |
|  | Anxiety | 0.84 (0.77, 0.91) | 1.05 (0.84, 1.30) | 2.07 (1.42, 3.03) | **1.94 (1.32, 2.85)** | 2.04 (1.28, 3.26) | 1.59 (0.99, 2.54) |
|  | Depression | 1.40 (1.31, 1.49) | 1.07 (0.87, 1.32) | 2.04 (1.45, 2.88) | **1.85 (1.31, 2.62)** | 1.04 (0.57, 1.89) | 0.91 (0.50, 1.65) |
|  | Pain | 1.14 (1.11, 1.17) | 1.01 (0.93, 1.09) | 1.19 (1.02, 1.39) | 1.02 (0.87, 1.20) | 1.21 (1.00, 1.45) | 1.03 (0.86, 1.25) |
| Non-CVD admission | SOB | 0.97 (0.96, 0.99) | 0.94 (0.91, 0.98) | 1.74 (1.62, 1.87) | **1.23 (1.14, 1.32)** | 1.77 (1.63, 1.92) | **1.21 (1.11, 1.32)** |
|  | Oedema | 1.18 (1.15, 1.20) | **1.07 (1.01, 1.13)** | 1.49 (1.34, 1.64) | **1.17 (1.06, 1.29)** | 1.61 (1.43, 1.81) | **1.25 (1.11, 1.41)** |
|  | Ankle swelling | 1.08 (1.04, 1.13) | 1.00 (0.91, 1.10) | 1.28 (1.04, 1.57) | 1.06 (0.86, 1.31) | 1.79 (1.44, 2.22) | **1.55 (1.24, 1.93)** |
|  | Fatigue | 1.17 (1.12, 1.22) | **1.12 (1.02, 1.24)** | 1.60 (1.36, 1.87) | **1.31 (1.12, 1.53)** | 1.67 (1.39, 2.00) | **1.32 (1.10, 1.58)** |
|  | Chest pain | 0.73 (0.67, 0.80) | 1.03 (0.97, 1.11) | 1.75 (1.56, 1.96) | 1.10 (0.98, 1.24) | 1.76 (1.55, 2.01) | 1.08 (0.94, 1.23) |
|  | Anxiety | 1.16 (1.10, 1.22) | 1.11 (0.97, 1.26) | 1.40 (1.13, 1.73) | 1.15 (0.93, 1.42) | 1.66 (1.33, 2.07) | 1.21 (0.97, 1.51) |
|  | Depression | 1.56 (1.48, 1.63) | **1.37 (1.22, 1.54)** | 1.72 (1.44, 2.04) | **1.46 (1.23, 1.74)** | 1.54 (1.25, 1.90) | **1.35 (1.10, 1.67)** |
|  | Pain | 1.21 (1.18, 1.23) | **1.15 (1.10, 1.21)** | 1.42 (1.33, 1.51) | **1.18 (1.10, 1.26)** | 1.46 (1.36, 1.57) | **1.23 (1.14, 1.33)** |
| Death | SOB | 0.72 (0.71, 0.73) | **0.76 (0.74, 0.79)** | 2.00 (1.80, 2.21) | **1.41 (1.27, 1.57)** | 1.93 (1.70, 2.19) | **1.25 (1.10, 1.43)** |
|  | Oedema | 1.04 (1.02, 1.06) | 0.97 (0.93, 1.02) | 1.94 (1.69, 2.23) | **1.37 (1.19, 1.58)** | 1.96 (1.65, 2.32) | **1.39 (1.17, 1.65)** |
|  | Ankle swelling | 0.98 (0.95, 1.02) | 1.00 (0.91, 1.09) | 1.98 (1.53, 2.58) | **1.49 (1.14, 1.94)** | 1.48 (1.02, 2.14) | 1.28 (0.88, 1.86) |
|  | Fatigue | 1.14 (1.10, 1.19) | 1.09 (0.99, 1.19) | 1.43 (1.11, 1.84) | 1.04 (0.80, 1.34) | 2.08 (1.62, 2.68) | **1.57 (1.22, 2.03)** |
|  | Chest pain | 0.60 (0.55, 0.65) | **0.79 (0.74, 0.85)** | 1.25 (1.02, 1.54) | 0.82 (0.66, 1.01) | 1.38 (1.10, 1.74) | 0.86 (0.68, 1.09) |
|  | Anxiety | 0.89 (0.85, 0.94) | 0.95 (0.83, 1.07) | 1.72 (1.28, 2.31) | **1.56 (1.16, 2.10)** | 0.95 (0.60, 1.49) | 0.69 (0.44, 1.09) |
|  | Depression | 1.27 (1.22, 1.33) | 1.22 (1.09, 1.36) | 1.66 (1.27, 2.17) | 1.47 (1.12, 1.93) | 1.62 (1.17, 2.23) | **1.48 (1.07, 2.04)** |
|  | Pain | 1.11 (1.09, 1.13) | 1.06 (1.01, 1.11) | 1.15 (1.03, 1.29) | 1.02 (0.91, 1.14) | 1.24 (1.16, 1.32) | 1.04 (0.91, 1.18) |

Flexible parametric models adjusted for age, sex, ethnicity, deprivation, BMI, diagnosis in hospital or primary care, previous hospitalisations comorbidities, and depression.

### **S4 Table:** **Complete-case analysis - Adjusted and unadjusted effect estimates of symptoms and outcomes at baseline, 6 and 12 month landmark times**

| Outcome | Symptom | Baseline | | 6 months | | 12 months | |
| --- | --- | --- | --- | --- | --- | --- | --- |
|  |  | **HR (95% CI)** | | **HR (95% CI)** | | **HR (95% CI)** | |
|  |  | Unadjusted | Adjusted | Unadjusted | Adjusted | Unadjusted | Adjusted |
| All-cause admission | SOB | 1.03 (1.00, 1.06) | 0.99 (0.95, 1.03) | 1.78 (1.67, 1.89) | 1.26 (1.17, 1.36) | 1.81 (1.68, 1.96) | 1.26 (1.14, 1.38) |
|  | Oedema | 1.10 (1.05, 1.15) | 1.01 (0.96, 1.07) | 1.49 (1.37, 1.63) | 1.16 (1.04, 1.30) | 1.66 (1.49, 1.86) | 1.26 (1.10, 1.44) |
|  | Ankle swelling | 1.02 (0.94, 1.10) | 0.99 (0.89, 1.09) | 1.33 (1.12, 1.60) | 1.03 (0.81, 1.29) | 1.64 (1.32, 2.04) | 1.45 (1.12, 1.87) |
|  | Fatigue | 1.16 (1.07, 1.26) | 1.09 (0.98, 1.21) | 1.49 (1.30, 1.72) | 1.26 (1.06, 1.50) | 1.83 (1.55, 2.16) | 1.39 (1.14, 1.70) |
|  | Chest pain | 1.18 (1.12, 1.24) | 1.10 (1.03, 1.18) | 1.83 (1.66, 2.02) | 1.12 (0.99, 1.28) | 1.83 (1.62, 2.08) | 1.11 (0.95, 1.30) |
|  | Anxiety | 1.13 (1.02, 1.26) | 1.05 (0.91, 1.21) | 1.47 (1.22, 1.76) | 1.18 (0.93, 1.50) | 1.58 (1.27, 1.97) | 1.25 (0.96, 1.63) |
|  | Depression | 1.37 (1.25, 1.51) | 1.05 (0.92, 1.20) | 1.68 (1.44, 1.96) | 1.17 (0.95, 1.43) | 1.56 (1.27, 1.91) | 1.28 (0.99, 1.64) |
|  | Pain | 1.17 (1.12, 1.21) | 1.09 (1.03, 1.14) | 1.32 (1.24, 1.40) | 1.11 (1.03, 1.19) | 1.35 (1.26, 1.45) | 1.14 (1.04, 1.24) |
| HF admission | SOB | 1.25 (1.18, 1.32) | 1.21 (1.12, 1.31) | 2.45 (2.14, 2.80) | 1.76 (1.48, 2.09) | 2.74 (2.29, 3.29) | 1.65 (1.30, 2.09) |
|  | Oedema | 1.25 (1.16, 1.36) | 1.04 (0.93, 1.15) | 1.97 (1.62, 2.39) | 1.48 (1.17, 1.88) | 2.25 (1.73, 2.92) | 1.66 (1.21, 2.27) |
|  | Ankle swelling | 1.13 (0.97, 1.31) | 0.98 (0.80, 1.19) | 1.73 (1.16, 2.57) | 1.20 (0.71, 2.05) | 1.31 (0.68, 2.52) | 1.36 (0.67, 2.74) |
|  | Fatigue | 1.24 (1.06, 1.45) | 1.10 (0.90, 1.35) | 1.79 (1.30, 2.47) | 1.43 (0.95, 2.15) | 2.41 (1.64, 3.54) | 1.83 (1.14, 2.94) |
|  | Chest pain | 1.04 (0.93, 1.15) | 0.91 (0.78, 1.06) | 1.57 (1.21, 2.03) | 0.76 (0.52, 1.09) | 1.66 (1.18, 2.35) | 0.97 (0.62, 1.51) |
|  | Anxiety | 1.07 (0.87, 1.33) | 1.20 (0.91, 1.57) | 2.07 (1.42, 3.03) | 1.83 (1.11, 3.02) | 2.11 (1.26, 3.51) | 2.27 (1.27, 4.06) |
|  | Depression | 1.12 (0.91, 1.38) | 0.94 (0.71, 1.26) | 2.04 (1.45, 2.88) | 1.84 (1.17, 2.88) | 1.17 (0.63, 2.18) | 1.42 (0.69, 2.90) |
|  | Pain | 1.05 (0.97, 1.14) | 0.99 (0.89, 1.10) | 1.19 (1.02, 1.39) | 0.95 (0.78, 1.16) | 1.12 (0.91, 1.38) | 0.87 (0.66, 1.14) |
| Non-CVD admission | SOB | 0.97 (0.93, 1.01) | 0.93 (0.89, 0.98) | 1.74 (1.62, 1.87) | 1.18 (1.08, 1.29) | 1.74 (1.59, 1.90) | 1.20 (1.08, 1.33) |
|  | Oedema | 1.17 (1.11, 1.23) | 1.04 (0.97, 1.11) | 1.49 (1.34, 1.64) | 1.11 (0.98, 1.26) | 1.65 (1.45, 1.87) | 1.27 (1.09, 1.47) |
|  | Ankle swelling | 1.03 (0.94, 1.13) | 0.99 (0.87, 1.12) | 1.28 (1.04, 1.57) | 0.96 (0.74, 1.26) | 1.80 (1.42, 2.29) | 1.52 (1.14, 2.01) |
|  | Fatigue | 1.19 (1.08, 1.32) | 1.16 (1.02, 1.31) | 1.60 (1.36, 1.87) | 1.37 (1.14, 1.66) | 1.66 (1.36, 2.02) | 1.19 (0.94, 1.52) |
|  | Chest pain | 1.07 (1.00, 1.14) | 1.07 (0.98, 1.17) | 1.75 (1.56, 1.96) | 1.15 (1.00, 1.33) | 1.74 (1.51, 2.01) | 1.10 (0.92, 1.31) |
|  | Anxiety | 1.25 (1.11, 1.42) | 1.10 (0.93, 1.30) | 1.40 (1.13, 1.73) | 1.13 (0.86, 1.48) | 1.39 (1.07, 1.81) | 0.98 (0.70, 1.36) |
|  | Depression | 1.51 (1.34, 1.69) | 1.08 (0.92, 1.26) | 1.72 (1.44, 2.04) | 1.11 (0.88, 1.40) | 1.53 (1.21, 1.93) | 1.20 (0.91, 1.60) |
|  | Pain | 1.26 (1.20, 1.32) | 1.15 (1.09, 1.23) | 1.42 (1.33, 1.51) | 1.18 (1.08, 1.28) | 1.46 (1.35, 1.58) | 1.24 (1.12, 1.36) |
| Death | SOB | 0.82 (0.78, 0.85) | 0.81 (0.77, 0.86) | 2.00 (1.81, 2.22) | 1.38 (1.20, 1.58) | 1.88 (1.63, 2.16) | 1.09 (0.90, 1.32) |
|  | Oedema | 1.15 (1.09, 1.22) | 0.95 (0.88, 1.02) | 1.94 (1.69, 2.23) | 1.35 (1.12, 1.62) | 1.92 (1.59, 2.33) | 1.30 (1.01, 1.66) |
|  | Ankle swelling | 1.06 (0.96, 1.17) | 0.96 (0.83, 1.11) | 2.00 (1.54, 2.61) | 1.33 (0.91, 1.96) | 1.16 (0.72, 1.86) | 1.13 (0.64, 2.00) |
|  | Fatigue | 1.25 (1.14, 1.38) | 1.15 (1.00, 1.32) | 1.40 (1.08, 1.81) | 1.11 (0.80, 1.53) | 1.91 (1.42, 2.56) | 1.52 (1.05, 2.19) |
|  | Chest pain | 0.81 (0.75, 0.88) | 0.81 (0.73, 0.91) | 1.27 (1.03, 1.56) | 0.84 (0.64, 1.10) | 1.36 (1.05, 1.76) | 0.86 (0.62, 1.21) |
|  | Anxiety | 1.06 (0.92, 1.22) | 1.09 (0.90, 1.32) | 1.66 (1.23, 2.25) | 1.17 (0.75, 1.81) | 0.98 (0.59, 1.62) | 0.60 (0.29, 1.28) |
|  | Depression | 1.36 (1.20, 1.53) | 1.12 (0.93, 1.35) | 1.68 (1.28, 2.20) | 1.22 (0.84, 1.77) | 1.49 (1.02, 2.18) | 1.51 (0.94, 2.44) |
|  | Pain | 1.09 (1.04, 1.15) | 1.05 (0.98, 1.13) | 1.14 (1.02, 1.28) | 0.94 (0.81, 1.09) | 1.14 (0.99, 1.31) | 1.03 (0.85, 1.23) |

Flexible parametric models adjusted for age, sex, ethnicity, deprivation, BMI, diagnosis in hospital or primary care, previous hospitalisations comorbidities, and depression.
